# Cost-effectiveness of osteoporotic fracture risk assessment in people with intellectual disabilities

**DOI:** 10.1101/2025.08.11.25333424

**Authors:** May Ee Png, Valeria Frighi, Tim A. Holt, Felix Achana, Margaret Smith, Gary S. Collins, Jan Roast, Stavros Petrou

**Author notes:** **Corresponding author** Tim Holt, Nuffield Department of Primary Care Health Sciences, University of Oxford, OX2 6GG, Oxford, UK.

## Abstract

**Background:** People with intellectual disabilities (ID) suffer higher rates of major osteoporotic fracture (MOF), including hip fracture (HF), at younger ages than the general population. We compared the cost-effectiveness of alternative fracture risk assessment strategies for people with ID aged ≥40 years from a UK National Health Services perspective over a lifetime horizon.

**Methods:** Three strategies were assessed: (S1) Risk assessment using the standard (QFracture) score at current policy thresholds; (S2) Use of a novel, tailored IDFracture risk score for all; and (S3) Conducting a one-time dual-energy X-ray absorptiometry (DXA) scan in all. S1 and S2 were followed by DXA scan for those at risk. At-risk individuals received recommended interventions. A decision-analytical model incorporated data from literature and national databases to calculate discounted direct healthcare costs, quality-adjusted life-years (QALYs), and incremental cost-effectiveness ratios (ICERs). Sensitivity and subgroup analyses were conducted.

**Results:** In the base-case, S2 (ICER: −£2,568/QALY) was dominant (i.e. less costly and more effective) and S3 (ICER: £1,678/QALY) was cost-effective relative to S1 for MOF. For HF, S2 (ICER: £32,116/QALY) and S3 (ICER: £49,536/QALY) were not cost-effective relative to S1 under the NICE-recommended cost-effectiveness thresholds. Findings from the sensitivity analyses were predominantly consistent with the base-case results. Subgroup analyses showed that age- and gender-specific strategies could be used.

**Conclusions:** For people with ID aged ≥40 years, a proactive approach to risk assessment for MOF is not only clinically beneficial, but also cost-effective.

**PLAIN ENGLISH SUMMARY:** People with intellectual disabilities (ID) are at higher risk of fracture, particularly hip fracture, due at least partly to thinning of the bones (osteoporosis). These fractures carry huge costs in human terms, and to the NHS. Finding better ways of preventing them is essential. In this study, we aimed to determine which of three risk assessment strategies is the most cost-effective at preventing fractures in these individuals.

The first strategy was the currently recommended approach, involving risk assessment in all women from age 65 and men from 75 years, or younger in those with a risk factor for osteoporosis. ID itself is not recognised as a risk factor in current guidelines. The second involved using IDFracture in people with ID at or above 40 years, followed by a bone density (DXA) scan for those found to be at risk. The third strategy involved a single DXA in those aged 40 years or over. In each strategy, preventive treatment would be offered if needed, based on the DXA result.

We found that the most cost-effective way of identifying people with risk above the intervention threshold of 10% over ten years for major osteoporotic fracture at age 40-79 years is to perform a DXA. The most cost-effective way of identifying people with risk above the intervention thresholds of 3% over ten years for hip fracture at age 40-79 years is to use QFracture and perform a DXA in those at risk. However, different strategies may be needed for different age and gender subgroups.

## INTRODUCTION

Based on a 2006 estimate of the global burden of osteoporotic fracture^1^, there were around nine million osteoporotic fractures annually (or one occurring every three seconds^2^). Osteoporotic fractures are associated with a reduction in quality of life, loss of functional independence, higher risk of mortality and higher economic costs^3^. This burden has undoubtedly grown over the years due to an ageing population and although not estimated, would be particularly significant among people with intellectual disabilities (ID), as the incidence of major osteoporotic fractures (MOF) and hip fractures (HF) are higher than the general population^4^.

Osteoporosis, a silent disease that may remain asymptomatic until fracture occurs, can be detected via a dual-energy x-ray absorptiometry (DXA), also known as the bone mineral density (BMD) scan, and treated with bone strengthening medicines. Current practice in the UK based on National Institute for Health and Clinical Excellence (NICE) and National Osteoporosis Guideline Group guidelines^5^, recommends use of a risk score known as ‘QFracture’ to be used to estimate the 10-year absolute risk of MOF and HF in the primary care setting. However, people with ID do not generally have their fracture risk assessed, and when they do, the standard risk scores are poorly tailored to this population. Their risk therefore goes unrecognised, and few are offered the opportunity to prevent fractures. A new tool (IDFracture) for assessing fracture risk in people with ID has been developed and validated (Smith et al, submitted for publication)^6^. IDFracture is an algorithm that generates a risk score estimating the 10-year risk of MOF and of HF for adults aged 30-79 years. Although recommended by NICE^5^, FRAX was not considered in this study because QFracture was favoured over FRAX in its more recent documentation on osteoporosis^7^.

We aimed to compare the cost-effectiveness of three potential fracture risk assessment strategies for MOF and HF among people with ID aged 40 years and above from the UK National Health Service (NHS) perspective over a lifetime horizon.

## METHODS

The results are reported in accordance with the Consolidated Health Economic Evaluation Reporting Standards 2022 (CHEERS 2022) statement^8^ for health economic evaluations.

### Fracture risk assessment strategies

The three fracture risk assessment strategies examined were:

(1) current practice based on guidance from NICE, entailing screening via QFracture^9^ for those deemed at risk by age and/or other criteria, followed by DXA for those above a treatment intervention threshold;^7,10^

(2) fracture risk assessment via IDFracture for all aged 40 years and above followed by DXA for those above a treatment intervention threshold; and

(3) DXA alone for all aged 40 years and above.

### Model structure

A decision tree with a Markov cohort model was constructed to reflect the options of the three fracture risk assessment strategies, in the primary care setting while modelling recurring outcomes through time to assess its costs and effects among those aged 40 years old from a UK NHS perspective over a lifetime horizon. The age of 40 years was chosen because our previous study^11^ indicated that this is the age at which people with ID would start to develop significant risk of osteoporotic fracture.

In the decision tree (Supplementary Fig. 1), based on the current NICE criteria for determining risk of MOF and HF using QFracture, we estimated that approximately 3% (1%-5%) of the population with intellectual disabilities would be screened at age 40-49 years. Those who tested positive (true and false positives) using the QFracture or IDFracture risk scores and were therefore considered at risk of fracture were treated according to their BMD T-scores after a diagnostic DXA. The treatment intervention threshold of testing positive for MOF was defined as at least 10% over ten years, and at least 3% over ten years for HF^5^. NICE does not offer specific guidance but advises clinicians to follow local protocols or other national guidelines for advice on intervention thresholds. Therefore, our choice was based on the most recent recommendations from the Scottish Intercollegiate Guideline Network, updated in 2021 for MOF^12^. For HF, our choice was based on the 2021 Clinical guideline for the prevention and treatment of osteoporosis by the National Osteoporosis Guideline Group^5^, which sets the intervention threshold at 2.3% and 3.5% ten-year risk at age 60 and 65 years respectively.

**Fig. 1.**
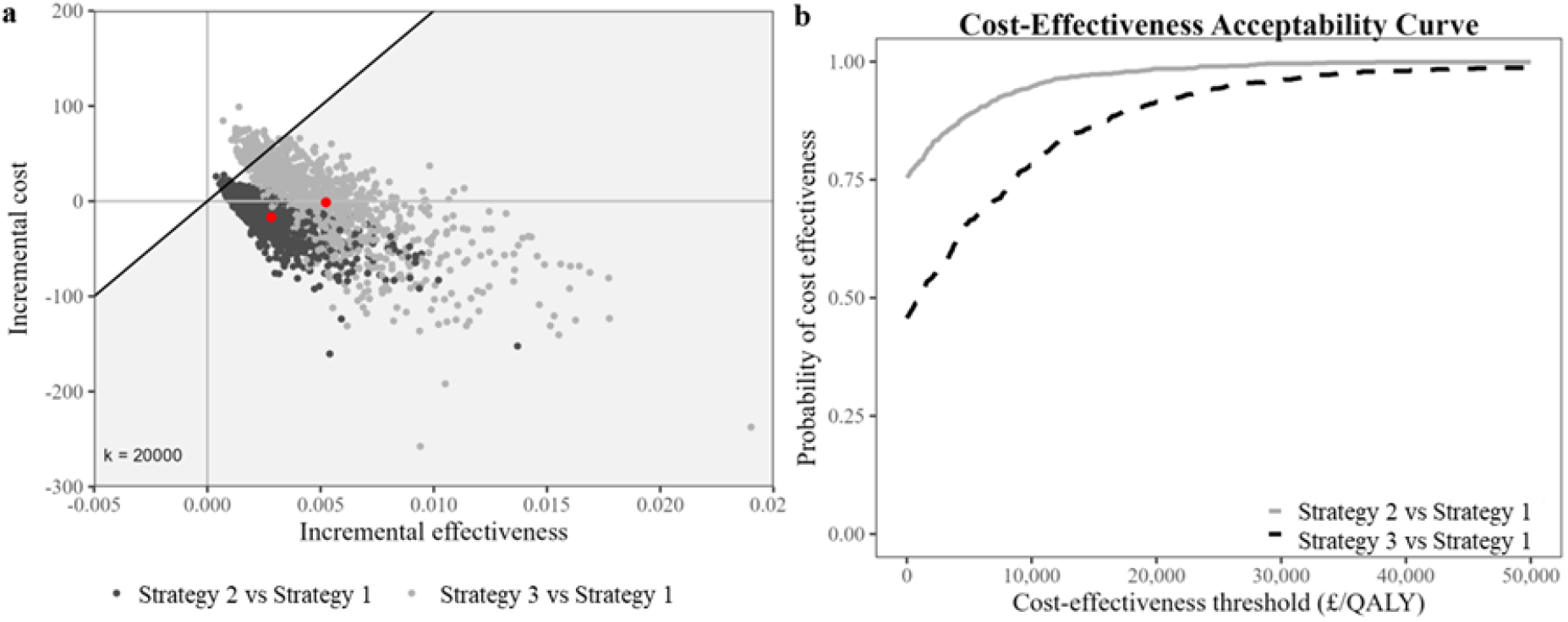
Cost-effectiveness analysis of fracture risk assessment strategies 1-3 among people with intellectual disabilities with major osteoporotic fracture using (a) cost-effectiveness plane and (b) cost-effectiveness acceptability curve.

The study cohort consisted of people aged 40-79 with ID, who were registered at their current practice at some point between 1/1/2008 and 31/10/2020 and were eligible for linkage to the Hospital Episode Statistics (HES)^13^ data and Index of Multiple deprivation. The HES dataset contains hospital records for the majority of patients attending secondary care services within the NHS in England. The diagnostic and service user codes used to identify people with ID has been described elsewhere^4^. Individuals above 79 years old were excluded because of the small number of individuals in this age group. We estimated 10-year risk of fracture by applying the IDFracture prediction models[6] to a cohort of people with ID, extracted from the Clinical Practice Research Datalink (CPRD) Aurum database. The CPRD Aurum captures data from over 40 million patients with 13 million current patients (20% of the UK population) over 1,000 general practices using EMIS Web® software^14,15^. We also applied a modified version of the QFracture 2012 algorithms^9^ to these data.

Based on the DXA, people who had a T-score of 2.5 standard deviation (SD) or more below the young adult mean were considered as having osteoporosis, those with a T-score that was between 1 and 2.5 SD below the young adult mean were considered as having osteopenia while those with BMD within 1 SD (+1 or −1) of the young adult mean were considered as having normal BMD^16^. Under the NICE guidance^5^, those with osteoporosis were treated using bisphosphonates in addition to vitamin D and calcium while those with osteopenia were treated with vitamin D and calcium alone. Since the NICE 2017 guidance noted that using the lowest available cost for each bisphosphonate in the cost-effectiveness modelling reflected clinical practice and would be appropriate^17^, alendronate 10 mg daily was selected among the bisphosphonates as the treatment of choice. We selected colecalciferol at a dose of 800 units and calcium at a dose of 1000 mg in view of the high prevalence of vitamin D deficiency in people with ID^18^ and of the standard calcium supplementation dose. All treatments were assumed to be given over a lifetime. No treatment was given to those with normal BMD.

The natural history of the condition and impact of treatment following fracture risk assessment was represented through a Markov model (Supplementary Fig. 2). This Markov model consisted of nine health states: untreated osteoporosis, treated osteoporosis, untreated osteopenia, treated osteopenia, normal BMD, no fracture, MOF or HF, post fracture and dead. The “population at risk” state is not a component of the Markov model but was presented to represent the movement of people in the decision tree. A person in the Markov model began in one of the osteopenia, osteoporosis, normal BMD or no fracture health states. Unless individuals were non-adherent to their treatment, they were assumed to not be able to move between “treated” and “untreated” health states. A separate “no fracture” health state was used to capture a proportion of the at-risk population (osteoporosis and osteopenia treated and untreated as well as people with normal BMD) who will not develop osteoporotic fracture in their lifetime.

**Fig. 2.**
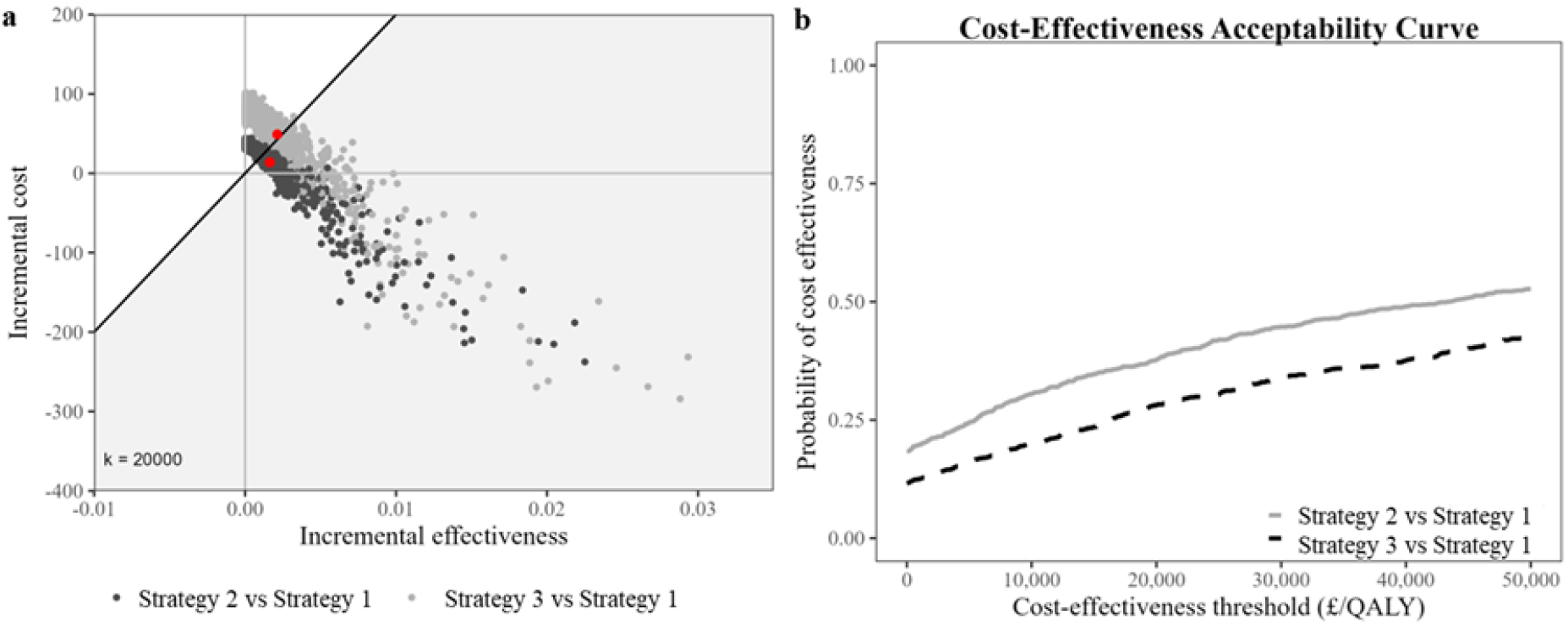
Cost-effectiveness analysis of fracture risk assessment strategies 1-3 among people with intellectual disabilities with hip fracture using (a) cost-effectiveness plane and (b) cost-effectiveness acceptability curve.

The model started with 1000 individuals and every year, each individual either remained in the same state or moved to another state based on the defined transition probabilities that were obtained from literature review; ‘death’ was the absorbing state (Supplementary Table 1). A one-year cycle length was assumed, and no half-cycle correction was applied, and the cohort was followed from 40 years old until an age of 100 years or death, whichever was earlier. The model was run separately for MOF and HF; ‘major osteoporotic fracture’ refers to a composite health state consisting of vertebral, shoulder, wrist, and hip fractures.

**Table 1.**
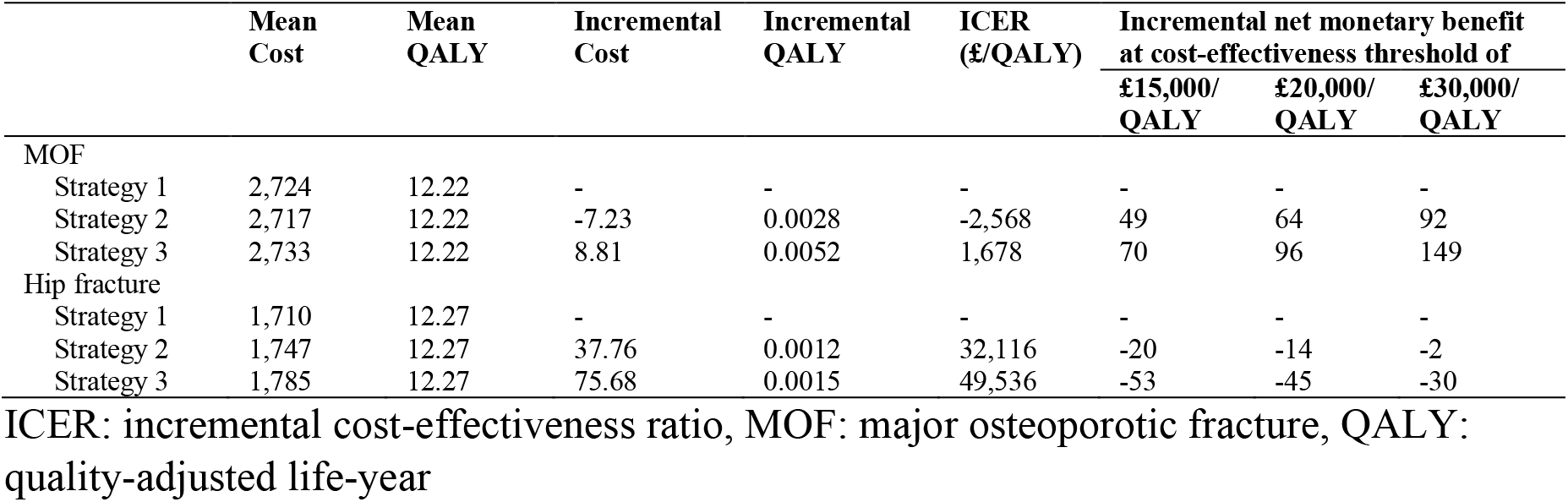
Base-case cost-effectiveness analysis of the three fracture risk assessment strategies.

### Model assumptions

There were a few assumptions made during the construct of the model.

- Non-osteoporotic fractures were excluded because other forms of fracture where the fracture risk assessment strategies would not be useful, should not be included in the modelling.
- Fracture risk assessment was assumed to occur once at the beginning of the model.
- DXA was assumed to be 100% accurate (i.e. have 100% sensitivity and specificity) in diagnosing osteoporosis and osteopenia.
- Side effects of bisphosphonates were excluded from the model since the risk of osteonecrosis related to bisphosphonate was very low; around 1 in 10,000 to 1 in 100,000 patient-years^19^.
- There is a 100% fracture risk assessment for Strategy 2.
- The post fracture cost is the same over time as the year 2 cost after fracture where year 1 is when the fracture occurred.

### Outcomes

Since utilities of people with ID were not available, we assumed that their utilities would be the same as the general population’s. These values were obtained from Janssen et al^20^, who used EuroQoL 5-Dimension (EQ-5D) index values based on UK-specific time trade-off value sets to obtain utility scores. Utility of treated osteoporosis, treated osteopenia, untreated osteoporosis and untreated osteopenia were assumed to be the same as the utility of a person who is fracture-free. The disutility values due to fracture and post-fractures were obtained from the International Costs and Utilities Related to Osteoporotic fractures Study (ICUROS) study^21^. Utility values between 0 (death) and 1 (full health) obtained were used to estimate the quality-adjusted life-years (QALYs) gained.

### Costs

The cost parameters are found in Supplementary Table 1. The unit cost of one DXA was obtained from the NHS Reference Costs^22^ while the unit cost of annual osteoporosis and osteopenia treatment was obtained from the Prescription Cost Analysis^23^.

Direct healthcare resource use and unit costs due to fracture and post-fracture comprised inpatient stays including operative procedures, accident and emergency attendances, outpatient visits as well as primary care visits and prescriptions and were obtained from the linked CPRD Aurum and HES databases of the same cohort of people aged 40-79 with ID.

An index MOF or HF among this cohort of people with ID was defined using a comprehensive list of primary care (SNOMED CT) terms and codes (for CPRD) and ICD-10 terms and codes (for HES) compiled by three clinicians; further details of this process have been described elsewhere^4^. Unit cost of hospital admission was obtained from the NHS Reference Cost 2021/22^22^ based on the clinical specialty, inpatient length of stay (short-stay versus long-stay) and type of admission (elective versus non-elective) after generating the Health Resource Group (HRG) codes from the HRG4+ 2021/22 Reference Cost Grouper at the Finished Consultant Episode (FCE) level^24,25^. Hospital admission costs were analysed at the episode level by summing FCE costs within episodes to generate total costs per inpatient episode. A similar process using this grouper was done to obtain the cost of emergency attendances and outpatient visits. The unit cost of primary care prescriptions was obtained from the estimates provided within the CPRD Aurum. All strategies included a general practitioner (GP) consultation cost. Unit cost of primary care consultations was obtained primarily from the Personal Social Services Research Unit (PSSRU) Unit Costs of Health and Social Care 2022 compendium^26^. All primary care consultations were assumed to last for 50 minutes, 9.22 minutes and 4 minutes for home, surgery and phone consultations respectively based on the average time reported in the PSSRU^26,27^. Staff below NHS Agenda for Change (AfC) pay band four were assumed to work 1618 hours per year^26^. Individuals who had died were assumed to incur zero service utilisation and costs.

Using a complete case analysis, the mean cost due to no fracture, fracture and post-fracture were computed using a two-parts model, which accounted for the skewed distribution of economic costs as a high frequency of patients incurred zero or low costs and a few patients incurred extremely high costs. This model had two stages - a logistic regression, in which the dependent variable (total cost) indicated presence of zero costs; followed by a generalised linear model with a Gaussian distribution and log-link function for economic costs relating to patients with positive values. The unit cost of post-fracture after the first-year post-fracture was assumed to be the same as the first-year post-fracture unit cost. The two-parts model was adjusted using age, gender, Index of Multiple Deprivation score and baseline cost (i.e. healthcare cost incurred within one year before the index fracture).

The adjusted mean total costs of fracture and post-fracture from the NHS perspective was computed from the sum of the adjusted mean costs of inpatient stays including operative procedures, emergency attendances, outpatient visits as well as primary care visits and prescriptions. All costs were adjusted to 2021/22 prices using the NHS Cost Inflation Index (NHSCII).^26^

### Base-case analysis

The cost-effectiveness of Strategy 2 (as previously described and applied to the whole population with ID aged 40-79 years), and Strategy 3 (as previously described and applied to the whole population with ID aged 40-79 years) were compared to Strategy 1 (as previously described and applied to 3% of the population with ID aged 40-79 years). Model outcomes were discounted QALYs and direct healthcare costs. The incremental cost-effectiveness ratio (ICER) was calculated as the incremental cost divided by the incremental QALYs and compared between the intervention strategy (i.e. Strategy 2 or Strategy 3) and the comparator strategy (i.e. Strategy 1). A discount rate of 3.5% was applied to outcomes and costs following the NICE guidelines^28^. The NICE cost-effectiveness thresholds of £20,000 per additional QALY^29^ and £30,000 per additional QALY^29^ were used to determine the cost-effectiveness of the fracture risk assessment strategies versus Strategy 1. An additional £15,000 per QALY cost-effectiveness threshold^30^ was also used to reflect recent trends in healthcare decision-making. A fracture risk assessment strategy with an ICER below these cost-effectiveness thresholds was considered to be cost-effective. The net monetary benefit (NMB) of the fracture risk assessment strategies was computed across different cost-effectiveness thresholds. A positive incremental NMB would indicate that the fracture risk assessment strategy is cost-effective compared with Strategy 1 at the given cost-effectiveness threshold. For completeness, the cost-effectiveness of Strategy 2 versus Strategy 3 was also compared. All analysis was conducted using R version 4.4^31^.

### Sensitivity analyses

Deterministic sensitivity analysis and probabilistic sensitivity analyses (PSA) were used to assess the impact of the parameter uncertainties on the cost-effectiveness results. For the former, different scenarios such as adherence to osteoporosis and osteopenia treatment, sensitivity and specificity of QFracture and IDFracture were varied from 20% less the base value to 100% while the costs listed in Supplementary Table 1 either halved or doubled from the base value. Fracture risk assessment using QFracture was varied between 1% and 5% according to expert opinion. As the fracture risk assessment in the model was over a 10-year period, but were followed-up over their lifetime to assess future benefits accrued, two sensitivity analyses were also performed based on a 10-year time horizon or a lifetime fracture risk, derived from the 10-year incidence rate using fixed effect meta-analysis. For the PSA, the parameters were sampled 1,000 times using beta distribution for probabilities and utilities and gamma distribution for costs. Simulations from the PSA were presented in a cost-effectiveness plane and a cost-effectiveness acceptability curve (CEAC).

### Subgroup analyses

Subgroup analyses stratified by age group and gender were performed. Based on current NICE criteria for determining risk of MOF and HF,^7,10^ we estimated that approximately 1% to 5% of the population with ID would be screened at age 40-49 years, approximately 35% to 50% at age 50-64 for women and 50-74 years for men, whilst all women and men with ID would qualify for screening from age 65 and 75 years respectively. Our estimates originated from the prevalence within the ID population of the factors considered by NICE as risk factors for MOF and HF and on the incidence of MOF and HF in our population.^4^

### Patient and public involvement

Our patient and public involvement representative, who is a co-investigator, helped shape the research question and advised on how best to share the findings, ensuring that communication was clear, realistic, and sensitive to patient concerns.

### Role of the funding source

The funder had no role in study design, data collection, data analysis, data interpretation, or writing of the report.

## RESULTS

As part of a check to ensure that the model works correctly, the predicted population size following screening at each health state during cycle 1 of the Markov model for both MOF and HF has been presented in Supplementary Table 2.

### Base-case analysis

For MOF, Strategy 2 (ICER: −£2,568/QALY) was dominant (i.e. less costly and more effective, on average) and Strategy 3 (ICER: £1,678/QALY) was cost-effective relative to Strategy 1 at the specified cost-effectiveness thresholds of £15,000/QALY to £30,000/QALY threshold (Table 1). On average, for MOF, Strategy 2 was £7.23 less costly and generated 0.0028 more QALYs than Strategy 1, and Strategy 3 was £8.81 more costly and generated 0.0052 more QALYs than Strategy 1. Strategies 2 and 3 had positive incremental NMB values relative to Strategy 1, thus indicating that they generate greater economic benefits than Strategy 1.

For HF, Strategy 2 (ICER: £32,116/QALY) and Strategy 3 (ICER: £49,536/QALY) were not cost-effective relative to Strategy 1 at the £15,000/QALY to £20,000/QALY thresholds (Table 1). On average, for HF, Strategy 2 was £37.76 more costly and generated 0.0012 more QALYs than Strategy 1, and Strategy 3 was £75.68 more costly and generated 0.0015 more QALYs than Strategy 1. Strategy 2 and Strategy 3 had negative incremental NMB values at the £15,000/QALY to £30,000/QALY thresholds, thus indicating that it would only be cost-effective relative to Strategy 1.

The comparison between Strategy 2 and Strategy 3 showed that showed that Strategy 3 was cost-effective relative to Strategy 2 (ICER: £6,594/QALY) for MOF (Supplementary Table 5) but it was not cost-effective relative to Strategy 2 (ICER: £107,731/QALY) for HF (Supplementary Table 6) at the specified cost-effectiveness thresholds.

### Deterministic sensitivity analyses

Findings from the sensitivity analyses for MOF (Supplementary Table 3) when Strategy 1 was the comparator were consistent with the base-case results except for Strategy 3 when compared to Strategy 1 under the £15,000/QALY threshold where the cost of DXA was doubled and a 10-year time horizon was applied; and under the £20,000/QALY threshold where a 10-year time horizon was applied.

Findings from the sensitivity analyses for HF (Supplementary Table 4) when Strategy 1 was the comparator were largely consistent with the base-case results. Exceptions where Strategy 2 became cost-effective relative to Strategy 1 at the £15,000/QALY threshold include when the adherence to osteoporosis treatment was increased to 100%; specificity of IDFracture was increased to 100%; and a lifetime fracture risk was adopted. Exceptions where Strategy 2 became cost-effective relative to Strategy 1 at the £20,000/QALY threshold include when the adherence to osteopenia treatment was increased to 100%; specificity of IDFracture increased to 100%; cost of DXA was doubled; cost of post-fracture was halved; and a lifetime fracture risk was adopted. Exceptions where Strategy 2 became cost-effective relative to Strategy 1 at the £30,000/QALY threshold include when the adherence to osteoporosis treatment was increased to 100%; sensitivity of IDFracture was increased to 99.9%; specificity of IDFracture was increased to 100%; cost of osteopenia treatment and DXA were halved; cost of fracture and post-fracture were doubled; and a lifetime fracture risk was adopted. Exceptions where Strategy 3 became cost-effective relative to Strategy 1 at the £15,000/QALY threshold include when a lifetime fracture risk was considered. Exceptions where Strategy 3 became cost-effective relative to Strategy 1 at the £20,000/QALY threshold include when cost of DXA was halved; and a lifetime fracture risk was adopted. Exceptions where Strategy 3 became cost-effective relative to Strategy 1 at the £30,000/QALY threshold include when the adherence to osteoporosis treatment was increased to 100%; cost of DXA was halved; and a lifetime fracture risk was adopted.

Findings from the sensitivity analyses for MOF (Supplementary Table 5) when Strategy 2 was compared with Strategy 3 were largely consistent with the base-case results except when the sensitivity of IDFracture for fracture was increased to 99· 9%; the cost of a DXA scan was doubled; and a 10-year time horizon was adopted under all the pre-specified cost-effectiveness thresholds. Further exceptions include when adherence to osteopenia treatment was increased to 100%; specificity of IDFracture was increased to 100%; and cost of post fracture was halved under the £15,000/QALY threshold.

Findings from the sensitivity analyses for HF (Supplementary Table 6) when Strategy 2 was compared with Strategy 3 were largely consistent with the base-case results except for when the cost of a DXA scan was halved under a £30,000/QALY threshold, and a lifetime fracture risk was considered under all the pre-specified cost-effectiveness thresholds.

### Probabilistic sensitivity analysis

In the cost-effectiveness plane for MOF (Fig. 1a), the scatter points of the PSA simulations were mainly distributed below the £20,000/QALY cost-effectiveness threshold for both Strategies 2 and 3. The CEAC (Fig. 1b) for both strategies showed an above 50% probability of being cost-effective at the NICE recommended cost-effectiveness thresholds. These suggest that both strategies are likely to be cost-effective relative to Strategy 1 at the specified cost-effectiveness thresholds.

In the cost-effectiveness plane for HF (Fig. 2a), the scatter points of the PSA simulations were mainly distributed above the £20,000/QALY threshold for Strategies 2 and 3. The CEAC (Fig. 2b) for Strategies 2 and 3 showed a below 50% probability of being cost-effective at the NICE recommended cost-effectiveness thresholds. These suggest that both strategies are not likely to be cost-effective relative to Strategy 1 at the pre-specified cost-effectiveness thresholds.

The scatter points of the PSA simulations were mainly distributed below the £20,000/QALY threshold when Strategy 3 was compared to Strategy 2 for MOF (Supplementary Fig. 1a). The CEAC showed an above 50% probability of being cost-effective at the NICE recommended cost-effectiveness thresholds (Supplementary Fig. 1b). This suggests that Strategy 3 is likely to be cost-effective relative to Strategy 2 at the specified cost-effectiveness thresholds.

The scatter points of the PSA simulations were mainly distributed above the £20,000/QALY threshold when Strategy 3 was compared to Strategy 2 for HF (Supplementary Fig. 2a). The CEAC showed a below 50% probability of being cost-effective at the NICE recommended cost-effectiveness thresholds (Supplementary Fig. 2b). This suggests that Strategy 3 is not likely to be cost-effective relative to Strategy 2 at the specified cost-effectiveness thresholds for HF.

### Subgroup analyses

Findings from the subgroup analyses for MOF (Supplementary Table 3) for Strategies 1 and 2, where the risk assessment threshold was stratified by age and gender, were the same as the base-case results except among men in general. Strategy 2 and Strategy 3 were no longer cost-effective relative to Strategy 1 under all the pre-specified cost-effectiveness thresholds for men aged 75-79 years. Further exception when Strategy 2 was no longer cost-effective relative to Strategy 1 was under the £15,000/QALY threshold among people aged 40-48 years and men aged 50-74 years for Strategy 1. Strategy 3 was no longer cost-effective relative to Strategy 1 under the £15,000/QALY to £20,000/QALY thresholds among people aged 40-49 years and men aged 50-74 years. Strategy 3 was also no longer cost-effective relative to Strategy 1 under the £30,000/QALY threshold among people aged 40-49 years and men aged 50-74 years at 50% risk assessment for Strategy 1.

Findings from the subgroup analyses for HF (Supplementary Table 4) mirrored the base-case results.

Findings from the subgroup analyses for MOF (Supplementary Table 5) comparing Strategies 2 and 3 mirrored the base-case results except among men aged 40-79 years and women aged 40-49 years.

Findings from the subgroup analyses for HF (Supplementary Table 6) comparing Strategies 2 and 3 mirrored the base-case results.

## DISCUSSION

This study showed that for people with ID aged 40-79 years, Strategy 3 (i.e. DXA alone) is more cost-effective than Strategy 2 (i.e. IDFracture followed by DXA) for MOF, and both would be more cost-effective than the current policy, Strategy 1 (i.e. QFracture followed by DXA in selected at risk groups). As HF is a subgroup of MOF, this finding is the most clinically important in terms of policy implication.

Looking in more detail, based on the estimates and assumptions of our health economics model, different strategies could be adopted for different age groups and genders. For MOF, DXA would be the most cost-effective among women with ID except among those aged 40-49 years where IDFracture followed by DXA would be the most cost-effective strategy. IDFracture followed by DXA would be the most cost-effective strategy among men with ID aged 40-74 years and QFracture followed by DXA among men with ID aged 75-79 years for MOF. For HF, QFracture followed by DXA would be the most cost-effective among the subgroups.

However, considering the population as a whole, DXA would be the most cost-effective strategy for MOF, and QFracture followed by DXA would be the most cost-effective strategy for HF among people with ID aged 40 years and above. We were interested in studying HF as an important outcome given its high relative risk, particularly in the younger people with ID, but HF is a subgroup of MOF and the MOF findings are primary in terms of clinical implication.

To the best of our knowledge, this is the first study that evaluated the cost-effectiveness of different fracture risk assessment strategies and certainly the first to do so in the ID community. The model we have constructed is different from published osteoporosis models^32–34^ because we chose to compare the costs and effects of fracture prevention in the osteoporosis and osteopenia health states separately. Our model compared three strategies, rather than the risk assessment tools that support them (QFracture, IDFracture, and DXA), in order to best inform future policy in addressing this area of health care.

However, this study is not without its limitations. First, many parameters are not based on people with ID but we have tried to use intellectual disability-specific estimates whenever available. One such example is the cost of fractures. Second, we did not include costs from the societal perspective, and this underestimates the additional economic burden of fractures (and the benefits of preventing them) because residential care or supportive living accommodation and individual productivity loss may be cost drivers^35^. We assumed that all people offered DXA scan would receive this investigation, but in fact a proportion of the ID community may not easily tolerate this procedure. Research to investigate more acceptable forms of measurement in this population is a priority. A proportion of people suffering an osteoporotic fracture may have had a previously normal DXA scan, a fact built into our model, and this sets a limit on the effectiveness of any prevention strategy based on BMD measurement. We had also assumed treatment over a lifetime due to model constraints, which is not the current practice where the need of continued treatment of bisphosphonate is re-evaluated after five years of bisphosphonate treatment^5^. This would overestimate the cost of osteoporosis treatment. We extrapolated the 10-year fracture risk to a lifetime time horizon linearly in our sensitivity analysis, but the fracture risk is likely to increase exponentially over time^4^. We had used the 2012 version of the QFracture algorithm, so our computation differed slightly from the latest 2016 version, and we did not have all variables used in the algorithm in our data e.g. family history of osteoporosis, living in a nursing or care home, so our QFracture predicted risks were not identical to those that would have been produced by the risk calculator. Last, we had assumed a constant post-fracture cost even though it is likely to decrease over time^36^.

## CONCLUSION

In conclusion, we have shown that current osteoporosis guidelines are not fit for purpose in people with ID, and that there is a pressing need to update them to prevent osteoporotic fractures, devastating for the individual and costly for the health system, in this high-risk population. A diagnosis of ID should be recognised not only as a risk factor (evidenced in previous studies), but also as an opportunity for cost-effective (in fact cost-saving) interventions to prevent fractures in this vulnerable group. This requires a systematic approach for those aged 40-79 years, using either a measurement of bone mineral density, or a risk assessment tool tailored to this population.

## Data availability

Electronic health records are, by definition, considered “sensitive” data in the UK by the Data Protection Act and cannot be shared via public deposition because of information governance restrictions in place to protect patient confidentiality. Access to data is possible only once approval has been obtained through the individual constituent entities controlling access to the data. The primary care data can be requested via application to the Clinical Practice Research Datalink (www.cprd.com/researcher), secondary care data can be requested via application to the hospital episode statistics from the UK Health and Social Care Information Centre (www.hscic.gov.uk/hesdata). The HES data used in this analysis are re-used with permission from NHS Digital who retain the copyright for that data.

## Acknowledgements

We first of all acknowledge the leadership and vision of Dr Valeria Frighi, the grant holder, who died tragically in March 2024, following completion of the main work presented here. Without her inspiration and tireless work this study would never have happened. We also acknowledge the main charities relevant to this research, namely the Downs Syndrome Association, the Royal Osteoporosis Society, and Mencap, who have been extremely supportive of this work in recent years.

This study is based in part on data from the Clinical Practice Research Datalink obtained under licence from the UK Medicines and Healthcare products Regulatory Agency. The data is provided by patients and collected by the NHS as part of their care and support. The interpretation and conclusions contained in this study are those of the author/s alone.

## Award information

This study was funded by the National Institute for Health Research Translating Research into Policy (NIHR TRiP, reference number: NIHR202094). The funder of the study had no role in study design, data collection, data analysis, data interpretation, or writing of the report.

## Conflicts of interest

VF, MS, GSC, and TAH report other grants from the National Institute for Health and Care Research (NIHR) during the conduct of this study. VF, MS and TAH also report grants from The Baily Thomas Charitable Fund. JR is the mother of an individual with intellectual disability.

## Ethical approval

The Clinical Practice Research Datalink (CPRD) has broad National Research Ethics Service approval to cover observational research using anonymised primary care data and established linkages. The study obtained ethics approval through CPRD Research Data Governance process (Protocol Number 21_000433 _).

