## Supplementary material for "Cost-effectiveness of osteoporotic fracture risk assessment in people with intellectual disabilities"

### TABLES

**Supplementary Table 1. Model parameters**

| Parameter | Value | Reference |
|---|---|---|
| **Transition probability** | | |
| Sensitivity of QFracture at 10% cutoff for MOF among people with ID (%) | 17.9 (95%CI 16.2-19.5) | HES-CPRD |
| Sensitivity of QFracture at 3% cutoff for hip fracture among people with ID (%) | 34.3 (95%CI 31.2-37.4) | HES-CPRD |
| Specificity of QFracture at 10% cutoff for MOF among people with ID (%) | 92.9 (9%%CI 92.6-93.1) | HES-CPRD |
| Specificity of QFracture at 3% cutoff for HF among people with ID (%) | 88.5 (95%CI 88.2-88.8) | HES-CPRD |
| Sensitivity of IDFracture at 10% cutoff for MOF among people with ID (%) | 53.9 (95%CI 51.7-56.0) | HES-CPRD |
| Sensitivity of IDFracture at 3% cutoff for HF among people with ID (%) | 77.2 (95%CI 74.4-79.9) | HES-CPRD |
| Specificity of IDFracture at 10% cutoff for MOF among people with ID (%) | 73.7 (95%CI 73.2-74.1) | HES-CPRD |
| Specificity of IDFracture at 3% cutoff for HF among people with ID (%) | 62.7 (95%CI 62.2-63.2) | HES-CPRD |
| Sensitivity of DXA (%) | 100 | assumed |
| Specificity of DXA (%) | 100 | assumed |
| Prevalence of osteopenia per year (%) | 33.2 | [1] |
| Prevalence of osteoporosis per year (%) | 41 | [1] |
| Probability of developing MOF within 10 years among people with ID | Men, 40-79yo: 0.0695 (95%CI 0.0650-0.0743)<br>Men, 40-49yo: 0.0442 (95%CI 0.0391-0.0500)<br>Men, 50-74yo: 0.0874 (95%CI 0.0804-0.0951)<br>Men, 75-79yo: 0.2432 (95%CI 0.1856-0.3149)<br><br>Women, 40-79yo: 0.1173 (95%CI 0.1106-0.1243)<br>Women, 40-49yo: 0.0606 (95%CI 0.0534-0.0687)<br>Women, 50-64yo: 0.1347 (95%CI 0.1236-0.1467)<br>Women, 65-79yo: 0.2565 (95%CI 0.2308-0.2845) | HES-CPRD |
| Probability of developing HF within 10 years among people with ID | Men, 40-79yo: 0.0352 (95%CI 0.0320-0.0387)<br>Men, 40-49yo: 0.0177 (95%CI 0.0145-0.0216)<br>Men, 50-74yo: 0.0476 (95%CI 0.0424-0.0534)<br>Men, 75-79yo: 0.1612 (95%CI 0.1134-0.2265)<br><br>Women, 40-79yo: 0.0456 (95%CI 0.0414-0.0503) | HES-CPRD |

| Parameter | Value | Reference |
|---|---|---|
| | Women, 40-49yo: 0.0127 (95%CI 0.0095-0.0168) | |
| | Women, 50-64yo: 0.0490 (95%CI 0.0421-0.0570) | |
| | Women, 65-79yo: 0.1509 (95%CI 0.1298-0.1750) | |
| Probability of developing MOF over a lifetime among people with ID | Men, 40-79yo: 0.1761 | derived from 10-year probability of developing MOF |
| | Men, 40-49yo: 0.0988 | |
| | Men, 50-74yo: 0.2014 | |
| | Men, 75-79yo: 0.4008 | |
| | Women, 40-79yo: 0.2902 | |
| | Women, 40-49yo: 0.1003 | |
| | Women, 50-64yo: 0.2732 | |
| | Women, 65-79yo: 0.4670 | |
| Probability of developing HF over a lifetime among people with ID | Men, 40-79yo: 0.0997 | derived from 10-year probability of developing HF |
| | Men, 40-49yo: 0.0378 | |
| | Men, 50-74yo: 0.1157 | |
| | Men, 75-79yo: 0.2476 | |
| | Women, 40-79yo: 0.1302 | |
| | Women, 40-49yo: 0.0218 | |
| | Women, 50-64yo: 0.1009 | |
| | Women, 65-79yo: 0.2401 | |
| Risk assessment (%) | 40-49 yo: 3 (range: 1-5)<br>Men, 50-74 yo: 35-50<br>Men, 75-79 yo: 100<br>Women, 50-64 yo: 35<br>Women, 65-79 yo: 100 | expert opinion |
| 10-year probability of MOF among people without ID but with a previous fragility fracture (%)* | Men: 9.6<br>Women: 16.4 | [2] |
| 10-year probability of HF among people without ID but with a previous fragility fracture (%)* | Men: 1.9<br>Women: 3.2 | [2] |
| Proportion of MOF that has osteoporosis (%) | Men: 20.69<br>Women: 44.09 | [3] |
| Proportion of HF that has osteoporosis (%) | Men: 38.89<br>Women: 63.79 | [3] |
| Proportion of MOF that has osteopenia (%) | Men: 61.38<br>Women: 43.29 | [3] |
| Proportion of HF that has osteopenia (%) | Men: 58.33<br>Women: 31.03 | [3] |
| Relative risk of MOF after alendronate** | 0.53 (95%CI 0.44-0.65) | [4] |
| Relative risk of HF after alendronate** | 0.61 (95%CI 0.39-0.95) | [4] |
| Relative risk of MOF after calcium and Vitamin D | 0.94 (95%CI 0.89-0.99) | [5] |
| Relative risk of HF after calcium and Vitamin D | 0.84 (95%CI 0.72-0.97) | [5] |
| Hazard ratio of death after MOF | Men: 2.00 (95%CI 1.88-2.12)<br>Women: 1.37 (95%CI 1.31-1.43) | [6] |
| Hazard ratio of death after HF | Men: 2.13 (95%CI 1.96-2.31)<br>Women: 1.60 (95%CI 1.50-1.70) | [6] |
| Adherence to osteoporosis treatment (%) | 43 (95%CI 38-49) | [7] |
| Adherence to osteopenia treatment among people with ID (%) | 80.4 | [8] |
| Mortality rate (per 1000 population) among people with ID | Men, 35-44yo: 4.4 (95%CI 2.8-6.7) | [9] |

| Parameter | Value | Reference |
|---|---|---|
| | Men, 45-54yo: 10.8 (95%CI 8.3-13.9) | |
| | Men, 55-64yo: 26.7 (95%CI 21.5-32.8) | |
| | Men, 65-74yo: 45.7 (95%CI 36.4-56.8) | |
| | Men, 75-84yo: 104.3 (95%CI 79.0-135.2) | |
| | Men, 85-99yo: 221.0 (95%CI 136.8-337.9) | |
| | Women, 35-44yo: 4.4 (95%CI 2.7-6.8) | |
| | Women, 45-54yo: 8.2 (95%CI 5.8-11.1) | |
| | Women, 55-64yo: 21.6 (95%CI 16.6-27.6) | |
| | Women, 65-74yo: 35.1 (95%CI 26.4-45.6) | |
| | Women, 75-84yo: 85.7 (95%CI 63.6-113.0) | |
| | Women, 85-99yo: 222.4 (95%CI 147.8-321.5) | |
| **Utility†** | | |
| Fracture-free | 35-44 yo: 0.893 | [10] |
| | 45-54 yo: 0.855 | |
| | 55-64 yo: 0.810 | |
| | 65-74 yo: 0.773 | |
| | 75+ yo: 0.703 | |
| **Disutility** | | |
| MOF | 0.27 (95%CI 0.24-0.29) | [11] |
| Hip fracture | 0.34 (95%CI 0.33-0.36) | [11] |
| Post MOF | 0.13 (95%CI 0.10-0.15) | [11] |
| Post hip fracture | 0.11 (95%CI 0.09-0.12) | [11] |
| **Annual cost (£), in 2021/22 prices** | | |
| DXA | 95.45 | [12] |
| General practitioner consultation | 41 | [13] |
| Bisphosphonate (i.e. Alendronate 10mg) | 91.25 | [14] |
| Calcium and vitamin D (i.e. Calcichew 1000mg/800unit) | 83.95 | [14] |
| First year MOF cost among people with ID‡ | 9,407 (SE 284) | HES-CPRD |
| First year HF cost among people with ID‡ | 12,761 (SE 410) | HES-CPRD |
| Post 1-year MOF cost among people with ID‡ | 5,687 (SE 594) | HES-CPRD |
| Post 1-year HF cost among people with ID‡ | 5,319 (SE 885) | HES-CPRD |

CI: confidence interval, CPRD: Clinical Practice Research Datalink, DXA: dual-energy x-ray absorptiometry, HES: Hospital Episode Statistics, HF: Hip fracture, HRQoL: health-related quality of life, ID: intellectual disabilities, MOF: major osteoporotic fracture, SE: standard error, yo: years old

*Converted to 1-year probability using the formula: $p' = 1-(1-p)^{1/t}$ where $p$ is the original probability and $t$ is time.

** People with alendronate in this network meta-analysis also had calcium and Vitamin D.

†Utility of treated osteoporosis, treated osteopenia, untreated osteoporosis and untreated osteopenia were assumed to be the same as the utility of a person who is fracture-free.

Recovery rate of major osteoporotic fracture was assumed to be the same as hip fracture.

‡Adjusted by age, gender, Indices of Multiple Deprivation quintile and baseline cost.

**Supplementary Table 2. Predicted population size following screening during cycle 1 of the Markov model for major osteoporotic fracture and hip fracture**

| | Untreated osteoporosis | Treated osteoporosis | Treated osteopenia | Untreated osteopenia | Normal BMD | No Fracture | Fracture | Post-fracture | Dead |
|---|---|---|---|---|---|---|---|---|---|
| Major osteoporotic fracture | | | | | | | | | |
| Strategy 1 | 38.08836122 | 0.2056388 | 0.1665173 | 30.84228274 | 24.0972 | 906.6 | 0 | 0 | 0 |
| Strategy 2 | 17.653534 | 20.64047 | 16.71374 | 14.2950568 | 24.0972 | 906.6 | 0 | 0 | 0 |
| Strategy 3 | 0 | 38.294 | 31.0088 | 0 | 24.0972 | 906.6 | 0 | 0 | 0 |
| Hip fracture | | | | | | | | | |
| Strategy 1 | 16.39355644 | 0.1704436 | 0.1380177 | 13.27478229 | 10.4232 | 959.6 | 0 | 0 | 0 |
| Strategy 2 | 3.776592 | 12.78741 | 10.35468 | 3.0581184 | 10.4232 | 959.6 | 0 | 0 | 0 |
| Strategy 3 | 0 | 16.564 | 13.4128 | 0 | 10.4232 | 959.6 | 0 | 0 | 0 |

BMD: bone mineral density, DXA: dual-energy x-ray absorptiometry

**Supplementary Table 3. Costs and effects of the fracture risk assessment strategies for major osteoporotic fracture based on sensitivity and subgroup analyses**

| | Mean Cost | Mean QALY | Incremental Cost | Incremental QALY | ICER (£/QALY) | Incremental net monetary benefit at cost-effectiveness threshold of | | |
|---|---|---|---|---|---|---|---|---|
| | | | | | | £15 000/ QALY | £20 000/ QALY | £30 000/ QALY |
| **Sensitivity analysis** | | | | | | | | |
| *Risk assessment at 1% for Strategy 1* | | | | | | | | |
| Strategy 1 | 2,724 | 12.22 | - | - | - | - | - | - |
| Strategy 2 | 2,717 | 12.22 | -7.32 | 0.0028 | -2,583 | 50 | 64 | 92 |
| Strategy 3 | 2,733 | 12.22 | 8.72 | 0.0053 | 1,655 | 70 | 97 | 149 |
| *Risk assessment at 5% for Strategy 1* | | | | | | | | |
| Strategy 1 | 2,724 | 12.22 | - | - | - | - | - | - |
| Strategy 2 | 2,717 | 12.22 | -7.14 | 0.0028 | -2,554 | 49 | 63 | 91 |
| Strategy 3 | 2,733 | 12.22 | 8.90 | 0.0052 | 1,701 | 70 | 96 | 148 |
| *Adherence to osteoporosis treatment -20%* | | | | | | | | |
| Strategy 1 | 2,744 | 12.21 | - | - | - | - | - | - |
| Strategy 2 | 2,737 | 12.22 | -6.62 | 0.0027 | -2,412 | 48 | 62 | 89 |
| Strategy 3 | 2,754 | 12.22 | 9.95 | 0.0051 | 1,946 | 67 | 92 | 143 |
| *Adherence to osteoporosis treatment at 100%* | | | | | | | | |
| Strategy 1 | 2,618 | 12.22 | - | - | - | - | - | - |
| Strategy 2 | 2,566 | 12.23 | -52.15 | 0.0074 | -7,007 | 164 | 201 | 275 |
| Strategy 3 | 2,543 | 12.24 | -74.91 | 0.0139 | -5,400 | 283 | 352 | 491 |
| *Adherence to osteopenia treatment -20%* | | | | | | | | |
| Strategy 1 | 2,762 | 12.21 | - | - | - | - | - | - |
| Strategy 2 | 2,750 | 12.22 | -11.77 | 0.0029 | -4,019 | 56 | 70 | 100 |
| Strategy 3 | 2,762 | 12.22 | 0.35 | 0.0055 | 63 | 82 | 109 | 163 |
| *Adherence to osteopenia treatment at 100%* | | | | | | | | |
| Strategy 1 | 2,684 | 12.22 | - | - | - | - | - | - |
| Strategy 2 | 2,696 | 12.22 | 11.64 | 0.0024 | 4,770 | 25 | 37 | 62 |
| Strategy 3 | 2,728 | 12.22 | 43.98 | 0.0045 | 9,672 | 24 | 47 | 92 |
| *Sensitivity of IDFracture for fracture -20%* | | | | | | | | |
| Strategy 1 | 2,724 | 12.22 | - | - | - | - | - | - |
| Strategy 2 | 2,725 | 12.22 | 0.75 | 0.0022 | 332 | 33 | 44 | 67 |
| Strategy 3 | 2,733 | 12.22 | 8.81 | 0.0052 | 1,678 | 70 | 96 | 149 |
| *Sensitivity of IDFracture for fracture at 99.9%** | | | | | | | | |
| Strategy 1 | 2,724 | 12.22 | - | - | - | - | - | - |
| Strategy 2 | 2,683 | 12.22 | -41.29 | 0.0052 | -7,873 | 120 | 146 | 199 |
| Strategy 3 | 2,733 | 12.22 | 8.81 | 0.0052 | 1,678 | 70 | 96 | 149 |
| *Specificity of IDFracture for fracture -20%* | | | | | | | | |
| Strategy 1 | 2,724 | 12.22 | - | - | - | - | - | - |
| Strategy 2 | 2,735 | 12.22 | 11.00 | 0.0028 | 3,906 | 31 | 45 | 73 |
| Strategy 3 | 2,733 | 12.22 | 8.81 | 0.0052 | 1,678 | 70 | 96 | 149 |
| *Specificity of IDFracture for fracture at 100%* | | | | | | | | |
| Strategy 1 | 2,724 | 12.22 | - | - | - | - | - | - |
| Strategy 2 | 2,684 | 12.22 | -39.77 | 0.0028 | -14,120 | 82 | 96 | 124 |
| Strategy 3 | 2,733 | 12.22 | 8.81 | 0.0052 | 1,678 | 70 | 96 | 149 |
| *Sensitivity of QFracture for fracture -20%* | | | | | | | | |
| Strategy 1 | 2,724 | 12.22 | - | - | - | - | - | - |

| | Mean Cost | Mean QALY | Incremental Cost | Incremental QALY | ICER (£/QALY) | Incremental net monetary benefit at cost-effectiveness threshold of | | |
|---|---|---|---|---|---|---|---|---|
| | | | | | | £15 000/ QALY | £20 000/ QALY | £30 000/ QALY |
| Strategy 2 | 2,717 | 12.22 | -7.31 | 0.0028 | -2,591 | 50 | 64 | 92 |
| Strategy 3 | 2,733 | 12.22 | 8.73 | 0.0053 | 1,661 | 70 | 96 | 149 |
| *Sensitivity of QFracture for fracture at 100%* | | | | | | | | |
| Strategy 1 | 2,722 | 12.22 | - | - | - | - | - | - |
| Strategy 2 | 2,717 | 12.22 | -5.41 | 0.0027 | -2,014 | 46 | 59 | 86 |
| Strategy 3 | 2,733 | 12.22 | 10.63 | 0.0051 | 2,077 | 66 | 92 | 143 |
| *Specificity of QFracture for fracture -20%* | | | | | | | | |
| Strategy 1 | 2,725 | 12.22 | - | - | - | - | - | - |
| Strategy 2 | 2,717 | 12.22 | -7.92 | 0.0028 | -2,813 | 50 | 64 | 92 |
| Strategy 3 | 2,733 | 12.22 | 8.12 | 0.0052 | 1,547 | 71 | 97 | 149 |
| *Specificity of QFracture for fracture at 100%* | | | | | | | | |
| Strategy 1 | 2,724 | 12.22 | - | - | - | - | - | - |
| Strategy 2 | 2,717 | 12.22 | -6.97 | 0.0028 | -2,475 | 49 | 63 | 91 |
| Strategy 3 | 2,733 | 12.22 | 9.07 | 0.0052 | 1,728 | 70 | 96 | 148 |
| *Cost of alendronate -50%* | | | | | | | | |
| Strategy 1 | 2,479 | 12.22 | - | - | - | - | - | - |
| Strategy 2 | 2,471 | 12.22 | -8.73 | 0.0028 | -3,101 | 51 | 65 | 93 |
| Strategy 3 | 2,485 | 12.22 | 6.01 | 0.0052 | 1,146 | 73 | 99 | 151 |
| *Cost of alendronate +100%* | | | | | | | | |
| Strategy 1 | 3,213 | 12.22 | - | - | - | - | - | - |
| Strategy 2 | 3,209 | 12.22 | -4.23 | 0.0028 | -1,503 | 46 | 61 | 89 |
| Strategy 3 | 3,228 | 12.22 | 14.40 | 0.0052 | 2,743 | 64 | 91 | 143 |
| *Cost of vitamin D and calcium -50%* | | | | | | | | |
| Strategy 1 | 2,317 | 12.22 | - | - | - | - | - | - |
| Strategy 2 | 2,306 | 12.22 | -11.22 | 0.0028 | -3,983 | 53 | 68 | 96 |
| Strategy 3 | 2,318 | 12.22 | 1.38 | 0.0052 | 263 | 77 | 104 | 156 |
| *Cost of vitamin D and calcium +100%* | | | | | | | | |
| Strategy 1 | 3,539 | 12.22 | - | - | - | - | - | - |
| Strategy 2 | 3,539 | 12.22 | 0.74 | 0.0028 | 261 | 42 | 56 | 84 |
| Strategy 3 | 3,562 | 12.22 | 23.66 | 0.0052 | 4,508 | 55 | 81 | 134 |
| *Cost of DXA -50%* | | | | | | | | |
| Strategy 1 | 2,724 | 12.22 | - | - | - | - | - | - |
| Strategy 2 | 2,703 | 12.22 | -20.90 | 0.0028 | -7,421 | 63 | 77 | 105 |
| Strategy 3 | 2,685 | 12.22 | -38.80 | 0.0052 | -7,391 | 118 | 144 | 196 |
| *Cost of DXA +100%* | | | | | | | | |
| Strategy 1 | 2,724 | 12.22 | - | - | - | - | - | - |
| Strategy 2 | 2,744 | 12.22 | 20.10 | 0.0028 | 7,136 | 22 | 36 | 64 |
| Strategy 3 | 2,828 | 12.22 | 104.03 | 0.0052 | 19,817 | -25 | 1 | 53 |
| *Cost of fracture -50%* | | | | | | | | |
| Strategy 1 | 2,600 | 12.22 | - | - | - | - | - | - |
| Strategy 2 | 2,597 | 12.22 | -3.14 | 0.0028 | -1,116 | 45 | 59 | 88 |
| Strategy 3 | 2,616 | 12.22 | 16.43 | 0.0052 | 3,130 | 62 | 89 | 141 |
| *Cost of fracture +100%* | | | | | | | | |
| Strategy 1 | 2,973 | 12.22 | - | - | - | - | - | - |

| | Mean Cost | Mean QALY | Incremental Cost | Incremental QALY | ICER (£/QALY) | Incremental net monetary benefit at cost-effectiveness threshold of | | |
|---|---|---|---|---|---|---|---|---|
| | | | | | | £15 000/ QALY | £20 000/ QALY | £30 000/ QALY |
| Strategy 2 | 2,957 | 12.22 | -15.41 | 0.0028 | -5,473 | 58 | 72 | 100 |
| Strategy 3 | 2,966 | 12.22 | -6.44 | 0.0052 | -1,226 | 85 | 111 | 164 |
| *Cost of post fracture -50%* | | | | | | | | |
| Strategy 1 | 2,159 | 12.22 | - | - | - | - | - | - |
| Strategy 2 | 2,176 | 12.22 | 17.31 | 0.0028 | 6,147 | 25 | 39 | 67 |
| Strategy 3 | 2,214 | 12.22 | 54.56 | 0.0052 | 10,394 | 24 | 50 | 103 |
| *Cost of post fracture +100%* | | | | | | | | |
| Strategy 1 | 3,854 | 12.22 | - | - | - | - | - | - |
| Strategy 2 | 3,798 | 12.22 | -56.33 | 0.0028 | -20,000 | 99 | 113 | 141 |
| Strategy 3 | 3,772 | 12.22 | -82.70 | 0.0052 | -15,754 | 161 | 188 | 240 |
| *10-year time horizon* | | | | | | | | |
| Strategy 1 | 1,146 | 7.17 | - | - | - | - | - | - |
| Strategy 2 | 1,160 | 7.17 | 13.78 | 0.0011 | 12,308 | 3 | 9 | 20 |
| Strategy 3 | 1,194 | 7.18 | 47.98 | 0.0021 | 22,990 | -17 | -6 | 15 |
| *Lifetime fracture risk* | | | | | | | | |
| Strategy 1 | 7,849 | 11.94 | - | - | - | - | - | - |
| Strategy 2 | 7,668 | 11.95 | -180.87 | 0.0120 | -15,047 | 361 | 421 | 541 |
| Strategy 3 | 7,525 | 11.96 | -324.58 | 0.0224 | -14,488 | 661 | 773 | 997 |
| **Subgroup analysis** | | | | | | | | |
| *40-49 years old* | | | | | | | | |
| Strategy 1 | 2,181 | 14.12 | - | - | - | - | - | - |
| Strategy 2 | 2,202 | 14.12 | 20.52 | 0.0010 | 19,603 | -5 | 0 | 11 |
| Strategy 3 | 2,245 | 14.13 | 63.40 | 0.0020 | 32,496 | -34 | -24 | -5 |
| *Men aged 50-74 years old, risk assessment at 35% for Strategy 1* | | | | | | | | |
| Strategy 1 | 1,568 | 9.74 | - | - | - | - | - | - |
| Strategy 2 | 1,592 | 9.74 | 23.70 | 0.0012 | 19,763 | -6 | 0 | 12 |
| Strategy 3 | 1,638 | 9.74 | 69.95 | 0.0024 | 29,643 | -35 | -23 | 1 |
| *Women aged 50-64 years old, risk assessment at 35% for Strategy 1* | | | | | | | | |
| Strategy 1 | 3,137 | 10.87 | - | - | - | - | - | - |
| Strategy 2 | 3,104 | 10.88 | -32.28 | 0.0030 | -10,708 | 77 | 93 | 123 |
| Strategy 3 | 3,093 | 10.88 | -43.51 | 0.0059 | -7,335 | 132 | 162 | 221 |
| *Men aged 50-74 years old, risk assessment at 50% for Strategy 1* | | | | | | | | |
| Strategy 1 | 1,569 | 9.74 | - | - | - | - | - | - |
| Strategy 2 | 1,592 | 9.74 | 22.72 | 0.0011 | 20,075 | -6 | 0 | 11 |
| Strategy 3 | 1,638 | 9.74 | 68.96 | 0.0023 | 30,088 | -35 | -23 | -0.2 |
| *Women aged 50-64 years old, risk assessment at 50% for Strategy 1* | | | | | | | | |
| Strategy 1 | 3,134 | 10.87 | - | - | - | - | - | - |
| Strategy 2 | 3,104 | 10.88 | -30.13 | 0.0028 | -10,591 | 73 | 87 | 115 |
| Strategy 3 | 3,093 | 10.88 | -41.36 | 0.0058 | -7,178 | 128 | 157 | 214 |
| *Men aged 75-79 years old, risk assessment at 100% for Strategy 1* | | | | | | | | |
| Strategy 1 | 773 | 5.07 | - | - | - | - | - | - |

| | Mean Cost | Mean QALY | Incremental Cost | Incremental QALY | ICER (£/QALY) | Incremental net monetary benefit at cost-effectiveness threshold of | | |
|---|---|---|---|---|---|---|---|---|
| | | | | | | **£15 000/ QALY** | **£20 000/ QALY** | **£30 000/ QALY** |
| Strategy 2 | 799 | 5.07 | 25.94 | 0.0005 | 52,222 | -18 | -16 | -11 |
| Strategy 3 | 854 | 5.07 | 80.52 | 0.0011 | 71,074 | -64 | -58 | -47 |
| *Women aged 65-79 years old, risk assessment at 100% for Strategy 1* | | | | | | | | |
| Strategy 1 | 1,691 | 6.95 | - | - | - | - | - | - |
| Strategy 2 | 1,688 | 6.96 | -2.59 | 0.0017 | -1,514 | 28 | 37 | 54 |
| Strategy 3 | 1,703 | 6.96 | 12.24 | 0.0039 | 3,135 | 46 | 66 | 105 |

DXA: dual x-ray absorptiometry, ICER: incremental cost-effectiveness ratio, QALY: quality-adjusted life-year
*99.9% instead of 100% was used as the high value for IDFracture sensitivity due to the incremental QALY between Strategy 2 and Strategy 3 to be less than <0.00001 when sensitivity of IDFracture is 100% and the ICER goes to infinity due to the small incremental QALY.

**Supplementary Table 4. Costs and effects of the fracture risk assessment strategies for hip fracture based on sensitivity and subgroup analyses**

| | Mean Cost | Mean QALY | Incremental Cost | Incremental QALY | ICER (£/QALY) | Incremental net monetary benefit at cost-effectiveness threshold of | | |
|---|---|---|---|---|---|---|---|---|
| | | | | | | £15 000/ QALY | £20 000/ QALY | £30 000/ QALY |
| **Sensitivity analysis** | | | | | | | | |
| *Risk assessment at 1% for Strategy 1* | | | | | | | | |
| Strategy 1 | 1,709 | 12.27 | - | - | - | - | - | - |
| Strategy 2 | 1,747 | 12.27 | 37.97 | 0.0012 | 32,003 | -20 | -14 | -2 |
| Strategy 3 | 1,785 | 12.27 | 75.89 | 0.0015 | 49,328 | -53 | -45 | -30 |
| *Risk assessment at 5% for Strategy 1* | | | | | | | | |
| Strategy 1 | 1,710 | 12.27 | - | - | - | - | - | - |
| Strategy 2 | 1,747 | 12.27 | 37.56 | 0.0012 | 32,232 | -20 | -14 | -3 |
| Strategy 3 | 1,785 | 12.27 | 75.48 | 0.0015 | 49,746 | -53 | -45 | -30 |
| *Adherence to osteoporosis treatment -20%* | | | | | | | | |
| Strategy 1 | 1,716 | 12.27 | - | - | - | - | - | - |
| Strategy 2 | 1,754 | 12.27 | 37.60 | 0.0012 | 32,467 | -20 | -14 | -3 |
| Strategy 3 | 1,792 | 12.27 | 75.47 | 0.0015 | 50,154 | -53 | -45 | -30 |
| *Adherence to osteoporosis treatment at 100%* | | | | | | | | |
| Strategy 1 | 1,675 | 12.27 | - | - | - | - | - | - |
| Strategy 2 | 1,712 | 12.28 | 37.15 | 0.0028 | 13,491 | 4 | 18 | 45 |
| Strategy 3 | 1,750 | 12.28 | 74.88 | 0.0036 | 20,930 | -21 | -3 | 32 |
| *Adherence to osteopenia treatment -20%* | | | | | | | | |
| Strategy 1 | 1,717 | 12.27 | - | - | - | - | - | - |
| Strategy 2 | 1,754 | 12.27 | 36.66 | 0.0012 | 31,615 | -19 | -13 | -2 |
| Strategy 3 | 1,791 | 12.27 | 74.25 | 0.0015 | 49,277 | -52 | -44 | -29 |
| *Adherence to osteopenia treatment at 100%* | | | | | | | | |
| Strategy 1 | 1,702 | 12.27 | - | - | - | - | - | - |
| Strategy 2 | 1,745 | 12.27 | 43.51 | 0.0013 | 32,804 | -24 | -17 | -4 |
| Strategy 3 | 1,785 | 12.27 | 83.14 | 0.0017 | 48,248 | -57 | -49 | -31 |
| *Sensitivity of IDFracture for fracture -20%* | | | | | | | | |
| Strategy 1 | 1,710 | 12.27 | - | - | - | - | - | - |
| Strategy 2 | 1,749 | 12.27 | 39.92 | 0.0009 | 42,578 | -26 | -21 | -12 |
| Strategy 3 | 1,785 | 12.27 | 75.68 | 0.0015 | 49,536 | -53 | -45 | -30 |
| *Sensitivity of IDFracture for fracture at 99.9%** | | | | | | | | |
| Strategy 1 | 1,710 | 12.27 | - | - | - | - | - | - |
| Strategy 2 | 1,744 | 12.27 | 34.60 | 0.0015 | 22,668 | -12 | -4 | 11 |
| Strategy 3 | 1,785 | 12.27 | 75.68 | 0.0015 | 49,536 | -53 | -45 | -30 |
| *Specificity of IDFracture for fracture -20%* | | | | | | | | |
| Strategy 1 | 1,710 | 12.27 | - | - | - | - | - | - |
| Strategy 2 | 1,764 | 12.27 | 54.18 | 0.0012 | 46,080 | -37 | -31 | -19 |
| Strategy 3 | 1,785 | 12.27 | 75.68 | 0.0015 | 49,536 | -53 | -45 | -30 |
| *Specificity of IDFracture for fracture at 100%* | | | | | | | | |
| Strategy 1 | 1,710 | 12.27 | - | - | - | - | - | - |
| Strategy 2 | 1,699 | 12.27 | -11.08 | 0.0012 | -9,419 | 29 | 35 | 46 |
| Strategy 3 | 1,785 | 12.27 | 75.68 | 0.0015 | 49,536 | -53 | -45 | -30 |
| *Sensitivity of QFracture for fracture -20%* | | | | | | | | |
| Strategy 1 | 1,710 | 12.27 | - | - | - | - | - | - |

| | Mean Cost | Mean QALY | Incremental Cost | Incremental QALY | ICER (£/QALY) | Incremental net monetary benefit at cost-effectiveness threshold of | | |
|---|---|---|---|---|---|---|---|---|
| | | | | | | £15 000/ QALY | £20 000/ QALY | £30 000/ QALY |
| Strategy 2 | 1,747 | 12.27 | 37.74 | 0.0012 | 32,005 | -20 | -14 | -2 |
| Strategy 3 | 1,785 | 12.27 | 75.65 | 0.0015 | 49,414 | -53 | -45 | -30 |
| *Sensitivity of QFracture for fracture at 100%* | | | | | | | | |
| Strategy 1 | 1,709 | 12.27 | - | - | - | - | - | - |
| Strategy 2 | 1,747 | 12.27 | 38.04 | 0.0011 | 33,209 | -21 | -15 | -4 |
| Strategy 3 | 1,785 | 12.27 | 75.96 | 0.0015 | 50,726 | -53 | -46 | -31 |
| *Specificity of QFracture for fracture -20%* | | | | | | | | |
| Strategy 1 | 1,710 | 12.27 | - | - | - | - | - | - |
| Strategy 2 | 1,747 | 12.27 | 37.07 | 0.0012 | 31,525 | -19 | -14 | -2 |
| Strategy 3 | 1,785 | 12.27 | 74.99 | 0.0015 | 49,080 | -52 | -44 | -29 |
| *Specificity of QFracture for fracture at 100%* | | | | | | | | |
| Strategy 1 | 1,709 | 12.27 | - | - | - | - | - | - |
| Strategy 2 | 1,747 | 12.27 | 38.22 | 0.0012 | 32,500 | -21 | -15 | -3 |
| Strategy 3 | 1,785 | 12.27 | 76.13 | 0.0015 | 49,831 | -53 | -46 | -30 |
| *Cost of alendronate -50%* | | | | | | | | |
| Strategy 1 | 1,451 | 12.27 | - | - | - | - | - | - |
| Strategy 2 | 1,487 | 12.27 | 36.84 | 0.0012 | 31,326 | -19 | -13 | -2 |
| Strategy 3 | 1,525 | 12.27 | 74.47 | 0.0015 | 48,746 | -52 | -44 | -29 |
| *Cost of alendronate +100%* | | | | | | | | |
| Strategy 1 | 2,227 | 12.27 | - | - | - | - | - | - |
| Strategy 2 | 2,267 | 12.27 | 39.62 | 0.0012 | 33,696 | -22 | -16 | -4 |
| Strategy 3 | 2,306 | 12.27 | 78.10 | 0.0015 | 51,115 | -55 | -48 | -32 |
| *Cost of vitamin D and calcium -50%* | | | | | | | | |
| Strategy 1 | 1,278 | 12.27 | - | - | - | - | - | - |
| Strategy 2 | 1,314 | 12.27 | 35.22 | 0.0012 | 29,953 | -18 | -12 | 0.1 |
| Strategy 3 | 1,351 | 12.27 | 72.38 | 0.0015 | 47,373 | -49 | -42 | -27 |
| *Cost of vitamin D and calcium +100%* | | | | | | | | |
| Strategy 1 | 2,572 | 12.27 | - | - | - | - | - | - |
| Strategy 2 | 2,615 | 12.27 | 42.85 | 0.0012 | 36,441 | -25 | -19 | -8 |
| Strategy 3 | 2,654 | 12.27 | 82.29 | 0.0015 | 53,861 | -59 | -52 | -36 |
| *Cost of DXA -50%* | | | | | | | | |
| Strategy 1 | 1,709 | 12.27 | - | - | - | - | - | - |
| Strategy 2 | 1,729 | 12.27 | 19.37 | 0.0012 | 16,474 | -2 | 4 | 16 |
| Strategy 3 | 1,738 | 12.27 | 28.13 | 0.0015 | 18,415 | -5 | 2 | 18 |
| *Cost of DXA +100%* | | | | | | | | |
| Strategy 1 | 1,710 | 12.27 | - | - | - | - | - | - |
| Strategy 2 | 1,784 | 12.27 | 74.55 | 0.0012 | 63,400 | -57 | -51 | -39 |
| Strategy 3 | 1,881 | 12.27 | 170.78 | 0.0015 | 111,777 | -148 | -140 | -125 |
| *Cost of fracture -50%* | | | | | | | | |
| Strategy 1 | 1,675 | 12.27 | - | - | - | - | - | - |
| Strategy 2 | 1,715 | 12.27 | 39.81 | 0.0012 | 33,853 | -22 | -16 | -5 |
| Strategy 3 | 1,754 | 12.27 | 78.33 | 0.0015 | 51,272 | -55 | -48 | -33 |
| *Cost of fracture +100%* | | | | | | | | |
| Strategy 1 | 1,778 | 12.27 | - | - | - | - | - | - |

| | Mean Cost | Mean QALY | Incremental Cost | Incremental QALY | ICER (£/QALY) | Incremental net monetary benefit at cost-effectiveness threshold of | | |
|---|---|---|---|---|---|---|---|---|
| | | | | | | £15 000/ QALY | £20 000/ QALY | £30 000/ QALY |
| Strategy 2 | 1,812 | 12.27 | 33.68 | 0.0012 | 28,643 | -16 | -10 | 2 |
| Strategy 3 | 1,848 | 12.27 | 70.37 | 0.0015 | 46,062 | -47 | -40 | -25 |
| *Cost of post fracture -50%* | | | | | | | | |
| Strategy 1 | 1,600 | 12.27 | - | - | - | - | - | - |
| Strategy 2 | 1,646 | 12.27 | 46.61 | 0.0012 | 39,635 | -29 | -23 | -11 |
| Strategy 3 | 1,687 | 12.27 | 87.17 | 0.0015 | 57,054 | -64 | -57 | -41 |
| *Cost of post fracture +100%* | | | | | | | | |
| Strategy 1 | 1,929 | 12.27 | - | - | - | - | - | - |
| Strategy 2 | 1,949 | 12.27 | 20.08 | 0.0012 | 17,078 | -2 | 3 | 15 |
| Strategy 3 | 1,982 | 12.27 | 52.71 | 0.0015 | 34,498 | -30 | -22 | -7 |
| *10-year time horizon* | | | | | | | | |
| Strategy 1 | 868 | 7.18 | - | - | - | - | - | - |
| Strategy 2 | 914 | 7.18 | 45.61 | 0.0004 | 108,412 | -39 | -37 | -33 |
| Strategy 3 | 954 | 7.18 | 85.88 | 0.0005 | 157,098 | -78 | -75 | -69 |
| *Lifetime fracture risk* | | | | | | | | |
| Strategy 1 | 3,240 | 12.18 | - | - | - | - | - | - |
| Strategy 2 | 3,179 | 12.19 | -60.76 | 0.0072 | -8,403 | 169 | 205 | 278 |
| Strategy 3 | 3,183 | 12.19 | -57.59 | 0.0094 | -6,130 | 199 | 246 | 339 |
| **Subgroup analysis** | | | | | | | | |
| *40-49 years old* | | | | | | | | |
| Strategy 1 | 1,697 | 14.15 | - | - | - | - | - | - |
| Strategy 2 | 1,747 | 14.15 | 49.97 | 0.0002 | 259,532 | -47 | -46 | -44 |
| Strategy 3 | 1,790 | 14.15 | 93.32 | 0.0003 | 373,010 | -90 | -88 | -86 |
| *Men aged 50-74 years old, risk assessment at 35% for Strategy 1* | | | | | | | | |
| Strategy 1 | 1,290 | 9.76 | - | - | - | - | - | - |
| Strategy 2 | 1,332 | 9.76 | 41.78 | 0.0007 | 59,658 | -31 | -28 | -21 |
| Strategy 3 | 1,373 | 9.76 | 82.61 | 0.0009 | 87,400 | -68 | -64 | -54 |
| *Women aged 50-64 years old, risk assessment at 35% for Strategy 1* | | | | | | | | |
| Strategy 1 | 1,620 | 10.93 | - | - | - | - | - | - |
| Strategy 2 | 1,651 | 10.94 | 31.07 | 0.0010 | 30,617 | -16 | -11 | -1 |
| Strategy 3 | 1,688 | 10.94 | 67.44 | 0.0014 | 49,231 | -47 | -40 | -26 |
| *Men aged 50-74 years old, risk assessment at 50% for Strategy 1* | | | | | | | | |
| Strategy 1 | 1,292 | 9.76 | - | - | - | - | - | - |
| Strategy 2 | 1,332 | 9.76 | 39.67 | 0.0006 | 61,497 | -30 | -27 | -20 |
| Strategy 3 | 1,373 | 9.76 | 80.50 | 0.0009 | 90,456 | -67 | -63 | -54 |
| *Women aged 50-64 years old, risk assessment at 50% for Strategy 1* | | | | | | | | |
| Strategy 1 | 1,621 | 10.93 | - | - | - | - | - | - |
| Strategy 2 | 1,651 | 10.94 | 29.79 | 0.0009 | 31,872 | -16 | -11 | -2 |
| Strategy 3 | 1,688 | 10.94 | 66.16 | 0.0013 | 51,297 | -47 | -40 | -27 |
| *Men aged 75-79 years old, risk assessment at 100% for Strategy 1* | | | | | | | | |
| Strategy 1 | 701 | 5.08 | - | - | - | - | - | - |
| Strategy 2 | 736 | 5.08 | 35.52 | 0.0003 | 140,028 | -32 | -30 | -28 |
| Strategy 3 | 779 | 5.08 | 77.89 | 0.0004 | 200,494 | -72 | -70 | -66 |

| | Mean Cost | Mean QALY | Incremental Cost | Incremental QALY | ICER (£/QALY) | Incremental net monetary benefit at cost-effectiveness threshold of | | |
|---|---|---|---|---|---|---|---|---|
| | | | | | | **£15 000/ QALY** | **£20 000/ QALY** | **£30 000/ QALY** |
| *Women aged 65-79 years old, risk assessment at 100% for Strategy 1* | | | | | | | | |
| Strategy 1 | 1,023 | 6.99 | - | - | - | - | - | - |
| Strategy 2 | 1,054 | 6.99 | 30.66 | 0.0005 | 61,866 | -23 | -21 | -16 |
| Strategy 3 | 1,093 | 6.99 | 69.76 | 0.0008 | 91,896 | -58 | -55 | -47 |

DXA: dual x-ray absorptiometry, ICER: incremental cost-effectiveness ratio, QALY: quality-adjusted life-year

*99.9% instead of 100% was used as the high value for IDFracture sensitivity due to the incremental QALY between Strategy 2 and Strategy 3 to be less than <0.00001 when sensitivity of IDFracture is 100% and the ICER goes to infinity due to the small incremental QALY.

**Supplementary Table 5. Cost-effectiveness analysis of Strategy 2 vs. Strategy 3 among people with intellectual disabilities with major osteoporotic fracture**

| | Mean Cost | Mean QALY | Incremental Cost | Incremental QALY | ICER (£/QALY) | Incremental net monetary benefit at cost-effectiveness threshold of | | |
|---|---|---|---|---|---|---|---|---|
| | | | | | | £15 000/ QALY | £20 000/ QALY | £30 000/ QALY |
| **Base case** | | | | | | | | |
| Strategy 2 | 2,717 | 12.22 | - | - | - | - | - | - |
| Strategy 3 | 2,733 | 12.22 | 16.04 | 0.0024 | 6,594 | 20 | 33 | 57 |
| **Sensitivity analysis** | | | | | | | | |
| *Adherence to osteoporosis treatment -20%* | | | | | | | | |
| Strategy 2 | 2,737 | 12.22 | - | - | - | - | - | - |
| Strategy 3 | 2,754 | 12.22 | 16.57 | 0.0024 | 6,991 | 19 | 31 | 55 |
| *Adherence to osteoporosis treatment at 100%* | | | | | | | | |
| Strategy 2 | 2,566 | 12.23 | - | - | - | - | - | - |
| Strategy 3 | 2,543 | 12.24 | -22.76 | 0.0064 | -3,540 | 119 | 151 | 216 |
| *Adherence to osteopenia treatment -20%* | | | | | | | | |
| Strategy 2 | 2,750 | 12.22 | - | - | - | - | - | - |
| Strategy 3 | 2,762 | 12.22 | 12.12 | 0.0025 | 4,789 | 26 | 39 | 64 |
| *Adherence to osteopenia treatment at 100%* | | | | | | | | |
| Strategy 2 | 2,696 | 12.22 | - | - | - | - | - | - |
| Strategy 3 | 2,728 | 12.22 | 32.35 | 0.0021 | 15,347 | -1 | 10 | 31 |
| *Sensitivity of IDFracture for fracture -20%* | | | | | | | | |
| Strategy 2 | 2,725 | 12.22 | - | - | - | - | - | - |
| Strategy 3 | 2,733 | 12.22 | 8.06 | 0.0030 | 2,686 | 37 | 52 | 82 |
| *Sensitivity of IDFracture for fracture at 99.9%** | | | | | | | | |
| Strategy 2 | 2,683 | 12.22 | - | - | - | - | - | - |
| Strategy 3 | 2,733 | 12.22 | 50.10 | <0.0001 | 9,491,917 | -50 | -50 | -50 |
| *Specificity of IDFracture for fracture -20%* | | | | | | | | |
| Strategy 2 | 2,735 | 12.22 | - | - | - | - | - | - |
| Strategy 3 | 2,733 | 12.22 | -2.19 | 0.0024 | -901 | 39 | 51 | 75 |
| *Specificity of IDFracture for fracture at 100%* | | | | | | | | |
| Strategy 2 | 2,684 | 12.22 | - | - | - | - | - | - |
| Strategy 3 | 2,733 | 12.22 | 48.58 | 0.0024 | 19,965 | -12 | 0 | 24 |
| *Cost of alendrontate -50%* | | | | | | | | |
| Strategy 2 | 2,471 | 12.22 | - | - | - | - | - | - |
| Strategy 3 | 2,485 | 12.22 | 14.75 | 0.0024 | 6,061 | 22 | 34 | 58 |
| *Cost of alendrontate +100%* | | | | | | | | |
| Strategy 2 | 3,209 | 12.22 | - | - | - | - | - | - |
| Strategy 3 | 3,228 | 12.22 | 18.64 | 0.0024 | 7,659 | 18 | 30 | 54 |
| *Cost of vitamin D and calcium treatment -50%* | | | | | | | | |
| Strategy 2 | 2,306 | 12.22 | - | - | - | - | - | - |
| Strategy 3 | 2,318 | 12.22 | 12.60 | 0.0024 | 5,179 | 24 | 36 | 60 |
| *Cost of vitamin D and calcium treatment +100%* | | | | | | | | |
| Strategy 2 | 3,539 | 12.22 | - | - | - | - | - | - |
| Strategy 3 | 3,562 | 12.22 | 22.93 | 0.0024 | 9,423 | 14 | 26 | 50 |
| *Cost of DXA -50%* | | | | | | | | |
| Strategy 2 | 2,703 | 12.22 | - | - | - | - | - | - |
| Strategy 3 | 2,685 | 12.22 | -17.90 | 0.0024 | -7,357 | 54 | 67 | 91 |

| | Mean Cost | Mean QALY | Incremental Cost | Incremental QALY | ICER (£/QALY) | Incremental net monetary benefit at cost-effectiveness threshold of | | |
|---|---|---|---|---|---|---|---|---|
| | | | | | | £15 000/ QALY | £20 000/ QALY | £30 000/ QALY |
| *Cost of DXA +100%* | | | | | | | | |
| Strategy 2 | 2,744 | 12.22 | - | - | - | - | - | - |
| Strategy 3 | 2,828 | 12.22 | 83.93 | 0.0024 | 34,495 | -47 | -35 | -11 |
| *Cost of fracture -50%* | | | | | | | | |
| Strategy 2 | 2,597 | 12.22 | - | - | - | - | - | - |
| Strategy 3 | 2,616 | 12.22 | 19.58 | 0.0024 | 8,046 | 17 | 29 | 53 |
| *Cost of fracture +100%* | | | | | | | | |
| Strategy 2 | 2,957 | 12.22 | - | - | - | - | - | - |
| Strategy 3 | 2,966 | 12.22 | 8.98 | 0.0024 | 3,689 | 28 | 40 | 64 |
| *Cost of post fracture -50%* | | | | | | | | |
| Strategy 2 | 2,176 | 12.22 | - | - | - | - | - | - |
| Strategy 3 | 2,214 | 12.22 | 37.25 | 0.0024 | 15,310 | -1 | 11 | 36 |
| *Cost of post fracture +100%* | | | | | | | | |
| Strategy 2 | 3,798 | 12.22 | - | - | - | - | - | - |
| Strategy 3 | 3,772 | 12.22 | -26.37 | 0.0024 | -10,838 | 63 | 75 | 99 |
| *10-year time horizon* | | | | | | | | |
| Strategy 2 | 1,160 | 7.17 | - | - | - | - | - | - |
| Strategy 3 | 1,194 | 7.18 | 34.20 | 0.0010 | 35,355 | -20 | -15 | -5 |
| *Lifetime fracture risk* | | | | | | | | |
| Strategy 2 | 7,668 | 11.95 | - | - | - | - | - | - |
| Strategy 3 | 7,525 | 11.96 | -143.71 | 0.0104 | -13,840 | 299 | 351 | 455 |
| **Subgroup analysis** | | | | | | | | |
| *40-49 years old* | | | | | | | | |
| Strategy 2 | 2,202 | 14.12 | - | - | - | - | - | - |
| Strategy 3 | 2,245 | 14.13 | 42.88 | 0.0009 | 47,420 | -29 | -25 | -16 |
| *Men aged 50-74 years old, risk assessment at 35% for Strategy 1* | | | | | | | | |
| Strategy 2 | 1,592 | 9.74 | - | - | - | - | - | - |
| Strategy 3 | 1,638 | 9.74 | 46.25 | 0.0012 | 39,851 | -29 | -23 | -11 |
| *Women aged 50-64 years old, risk assessment at 35% for Strategy 1* | | | | | | | | |
| Strategy 2 | 3,104 | 10.88 | - | - | - | - | - | - |
| Strategy 3 | 3,093 | 10.88 | -11.23 | 0.0029 | -3,850 | 55 | 70 | 99 |
| *Men aged 50-74 years old, risk assessment at 50% for Strategy 1* | | | | | | | | |
| Strategy 2 | 1,592 | 9.74 | - | - | - | - | - | - |
| Strategy 3 | 1,638 | 9.74 | 46.25 | 0.0012 | 39,851 | -29 | -23 | -11 |
| *Women aged 50-64 years old, risk assessment at 50% for Strategy 1* | | | | | | | | |
| Strategy 2 | 3,104 | 10.88 | - | - | - | - | - | - |
| Strategy 3 | 3,093 | 10.88 | -11.23 | 0.0029 | -3,850 | 55 | 70 | 99 |
| *Men aged 75-79 years old, risk assessment at 100% for Strategy 1* | | | | | | | | |
| Strategy 2 | 799 | 5.07 | - | - | - | - | - | - |
| Strategy 3 | 854 | 5.07 | 54.58 | 0.0006 | 85,795 | -45 | -42 | -35 |

| | Mean Cost | Mean QALY | Incremental Cost | Incremental QALY | ICER (£/QALY) | Incremental net monetary benefit at cost-effectiveness threshold of | | |
|---|---|---|---|---|---|---|---|---|
| | | | | | | £15 000/ QALY | £20 000/ QALY | £30 000/ QALY |
| *Women aged 65-79 years old, risk assessment at 100% for Strategy 1* | | | | | | | | |
| Strategy 2 | 1,688 | 6.96 | - | - | - | - | - | - |
| Strategy 3 | 1,703 | 6.96 | 14.83 | 0.0022 | 6,766 | 18 | 29 | 51 |

DXA: dual x-ray absorptiometry, ICER: incremental cost-effectiveness ratio, QALY: quality-adjusted life-year

*99.9% instead of 100% was used as the high value for IDFracture sensitivity due to the incremental QALY between Strategy 2 and Strategy 3 to be less than <0.00001 when sensitivity of IDFracture is 100% and the ICER goes to infinity due to the small incremental QALY.

### Supplementary Table 6. Cost-effectiveness analysis of Strategy 2 vs. Strategy 3 among people with intellectual disabilities with hip fracture

| | Mean Cost | Mean QALY | Incremental Cost | Incremental QALY | ICER (£/QALY) | Incremental net monetary benefit at cost-effectiveness threshold of | | |
|---|---|---|---|---|---|---|---|---|
| | | | | | | £15 000/ QALY | £20 000/ QALY | £30 000/ QALY |
| **Base case** | | | | | | | | |
| Strategy 2 | 1,747 | 12.27 | - | - | - | - | - | - |
| Strategy 3 | 1,785 | 12.27 | 37.92 | 0.0004 | 107,731 | -33 | -31 | -27 |
| **Sensitivity analysis** | | | | | | | | |
| *Adherence to osteoporosis treatment -20%* | | | | | | | | |
| Strategy 2 | 1,754 | 12.27 | - | - | - | - | - | - |
| Strategy 3 | 1,792 | 12.27 | 37.87 | 0.0003 | 109,245 | -33 | -31 | -27 |
| *Adherence to osteoporosis treatment at 100%* | | | | | | | | |
| Strategy 2 | 1,712 | 12.28 | - | - | - | - | - | - |
| Strategy 3 | 1,750 | 12.28 | 37.73 | 0.0008 | 45,782 | -25 | -21 | -13 |
| *Adherence to osteopenia treatment -20%* | | | | | | | | |
| Strategy 2 | 1,754 | 12.27 | - | - | - | - | - | - |
| Strategy 3 | 1,791 | 12.27 | 37.59 | 0.0003 | 108,285 | -32 | -31 | -27 |
| *Adherence to osteopenia treatment at 100%* | | | | | | | | |
| Strategy 2 | 1,745 | 12.27 | - | - | - | - | - | - |
| Strategy 3 | 1,785 | 12.27 | 39.64 | 0.0004 | 99,846 | -34 | -32 | -28 |
| *Sensitivity of IDFracture for fracture -20%* | | | | | | | | |
| Strategy 2 | 1,749 | 12.27 | - | - | - | - | - | - |
| Strategy 3 | 1,785 | 12.27 | 35.76 | 0.0006 | 60,585 | -27 | -24 | -18 |
| *Sensitivity of IDFracture for fracture at 99.9%** | | | | | | | | |
| Strategy 2 | 1,744 | 12.27 | - | - | - | - | - | - |
| Strategy 3 | 1,785 | 12.27 | 41.08 | <0.0001 | 26,613,676 | -41 | -41 | -41 |
| *Specificity of IDFracture for fracture -20%* | | | | | | | | |
| Strategy 2 | 1,764 | 12.27 | - | - | - | - | - | - |
| Strategy 3 | 1,785 | 12.27 | 21.50 | 0.0004 | 61,080 | -16 | -14 | -11 |
| *Specificity of IDFracture for fracture at 100%* | | | | | | | | |
| Strategy 2 | 1,699 | 12.27 | - | - | - | - | - | - |
| Strategy 3 | 1,785 | 12.27 | 86.76 | 0.0004 | 246,494 | -81 | -80 | -76 |
| *Cost of alendrontate -50%* | | | | | | | | |
| Strategy 2 | 1,487 | 12.27 | - | - | - | - | - | - |
| Strategy 3 | 1,525 | 12.27 | 37.64 | 0.0004 | 106,941 | -32 | -31 | -27 |
| *Cost of alendrontate +100%* | | | | | | | | |
| Strategy 2 | 2,267 | 12.27 | - | - | - | - | - | - |
| Strategy 3 | 2,306 | 12.27 | 38.47 | 0.0004 | 109,311 | -33 | -31 | -28 |
| *Cost of vitamin D and calcium treatment -50%* | | | | | | | | |
| Strategy 2 | 1,314 | 12.27 | - | - | - | - | - | - |
| Strategy 3 | 1,351 | 12.27 | 37.16 | 0.0004 | 105,568 | -32 | -30 | -27 |
| *Cost of vitamin D and calcium treatment +100%* | | | | | | | | |
| Strategy 2 | 2,615 | 12.27 | - | - | - | - | - | - |
| Strategy 3 | 2,654 | 12.27 | 39.44 | 0.0004 | 112,057 | -34 | -32 | -29 |
| *Cost of DXA -50%* | | | | | | | | |
| Strategy 2 | 1,729 | 12.27 | - | - | - | - | - | - |
| Strategy 3 | 1,738 | 12.27 | 8.76 | 0.0004 | 24,899 | -3 | -2 | 2 |

| | Mean Cost | Mean QALY | Incremental Cost | Incremental QALY | ICER (£/QALY) | Incremental net monetary benefit at cost-effectiveness threshold of | | |
|---|---|---|---|---|---|---|---|---|
| | | | | | | £15 000/ QALY | £20 000/ QALY | £30 000/ QALY |
| *Cost of DXA +100%* | | | | | | | | |
| Strategy 2 | 1,784 | 12.27 | - | - | - | - | - | - |
| Strategy 3 | 1,881 | 12.27 | 96.23 | 0.0004 | 273,396 | -91 | -89 | -86 |
| *Cost of fracture -50%* | | | | | | | | |
| Strategy 2 | 1,715 | 12.27 | - | - | - | - | - | - |
| Strategy 3 | 1,754 | 12.27 | 38.53 | 0.0004 | 109,468 | -33 | -31 | -28 |
| *Cost of fracture +100%* | | | | | | | | |
| Strategy 2 | 1,812 | 12.27 | - | - | - | - | - | - |
| Strategy 3 | 1,848 | 12.27 | 36.70 | 0.0004 | 104,258 | -31 | -30 | -26 |
| *Cost of post fracture -50%* | | | | | | | | |
| Strategy 2 | 1,646 | 12.27 | - | - | - | - | - | - |
| Strategy 3 | 1,687 | 12.27 | 40.56 | 0.0004 | 115,250 | -35 | -34 | -30 |
| *Cost of post fracture +100%* | | | | | | | | |
| Strategy 2 | 1,949 | 12.27 | - | - | - | - | - | - |
| Strategy 3 | 1,982 | 12.27 | 32.62 | 0.0004 | 92,693 | -27 | -26 | -22 |
| *10-year time horizon* | | | | | | | | |
| Strategy 2 | 914 | 7.18 | - | - | - | - | - | - |
| Strategy 3 | 954 | 7.18 | 40.27 | 0.0001 | 319,747 | -38 | -38 | -36 |
| *Lifetime fracture risk* | | | | | | | | |
| Strategy 2 | 3,179 | 12.19 | - | - | - | - | - | - |
| Strategy 3 | 3,183 | 12.19 | 3.17 | 0.0022 | 1,465 | 29 | 40 | 62 |
| **Subgroup analysis** | | | | | | | | |
| *40-49 years old* | | | | | | | | |
| Strategy 2 | 1,747 | 14.15 | - | - | - | - | - | - |
| Strategy 3 | 1,790 | 14.15 | 43.35 | <0.0001 | 752,122 | -42 | -42 | -42 |
| *Men aged 50-74 years old, risk assessment at 35% for Strategy 1* | | | | | | | | |
| Strategy 2 | 1,332 | 9.76 | - | - | - | - | - | - |
| Strategy 3 | 1,373 | 9.76 | 40.83 | 0.0002 | 166,726 | -37 | -36 | -33 |
| *Women aged 50-64 years old, risk assessment at 35% for Strategy 1* | | | | | | | | |
| Strategy 2 | 1,651 | 10.94 | - | - | - | - | - | - |
| Strategy 3 | 1,688 | 10.94 | 36.37 | 0.0004 | 102,457 | -31 | -29 | -26 |
| *Men aged 50-74 years old, risk assessment at 50% for Strategy 1* | | | | | | | | |
| Strategy 2 | 1,332 | 9.76 | - | - | - | - | - | - |
| Strategy 3 | 1,373 | 9.76 | 40.83 | 0.0002 | 166,726 | -37 | -36 | -33 |
| *Women aged 50-64 years old, risk assessment at 50% for Strategy 1* | | | | | | | | |
| Strategy 2 | 1,651 | 10.94 | - | - | - | - | - | - |
| Strategy 3 | 1,688 | 10.94 | 36.37 | 0.0004 | 102,457 | -31 | -29 | -26 |
| *Men aged 75-79 years old, risk assessment at 100% for Strategy 1* | | | | | | | | |
| Strategy 2 | 736 | 5.08 | - | - | - | - | - | - |
| Strategy 3 | 779 | 5.08 | 42.37 | 0.0001 | 314,266 | -40 | -40 | -38 |

| | Mean Cost | Mean QALY | Incremental Cost | Incremental QALY | ICER (£/QALY) | Incremental net monetary benefit at cost-effectiveness threshold of | | |
|---|---|---|---|---|---|---|---|---|
| | | | | | | **£15 000/ QALY** | **£20 000/ QALY** | **£30 000/ QALY** |
| *Women aged 65-79 years old, risk assessment at 100% for Strategy 1* | | | | | | | | |
| Strategy 2 | 1,054 | 6.99 | - | - | - | - | - | - |
| Strategy 3 | 1,093 | 6.99 | 39.09 | 0.0003 | 148,401 | -35 | -34 | -31 |

DXA: dual x-ray absorptiometry, ICER: incremental cost-effectiveness ratio, QALY: quality-adjusted life-year

*99.9% instead of 100% was used as the high value for IDFracture sensitivity due to the incremental QALY between Strategy 2 and Strategy 3 to be less than <0.00001 when sensitivity of IDFracture is 100% and the ICER goes to infinity due to the small incremental QALY.

**FIGURES**

**Supplementary Fig. 1. Decision tree model**

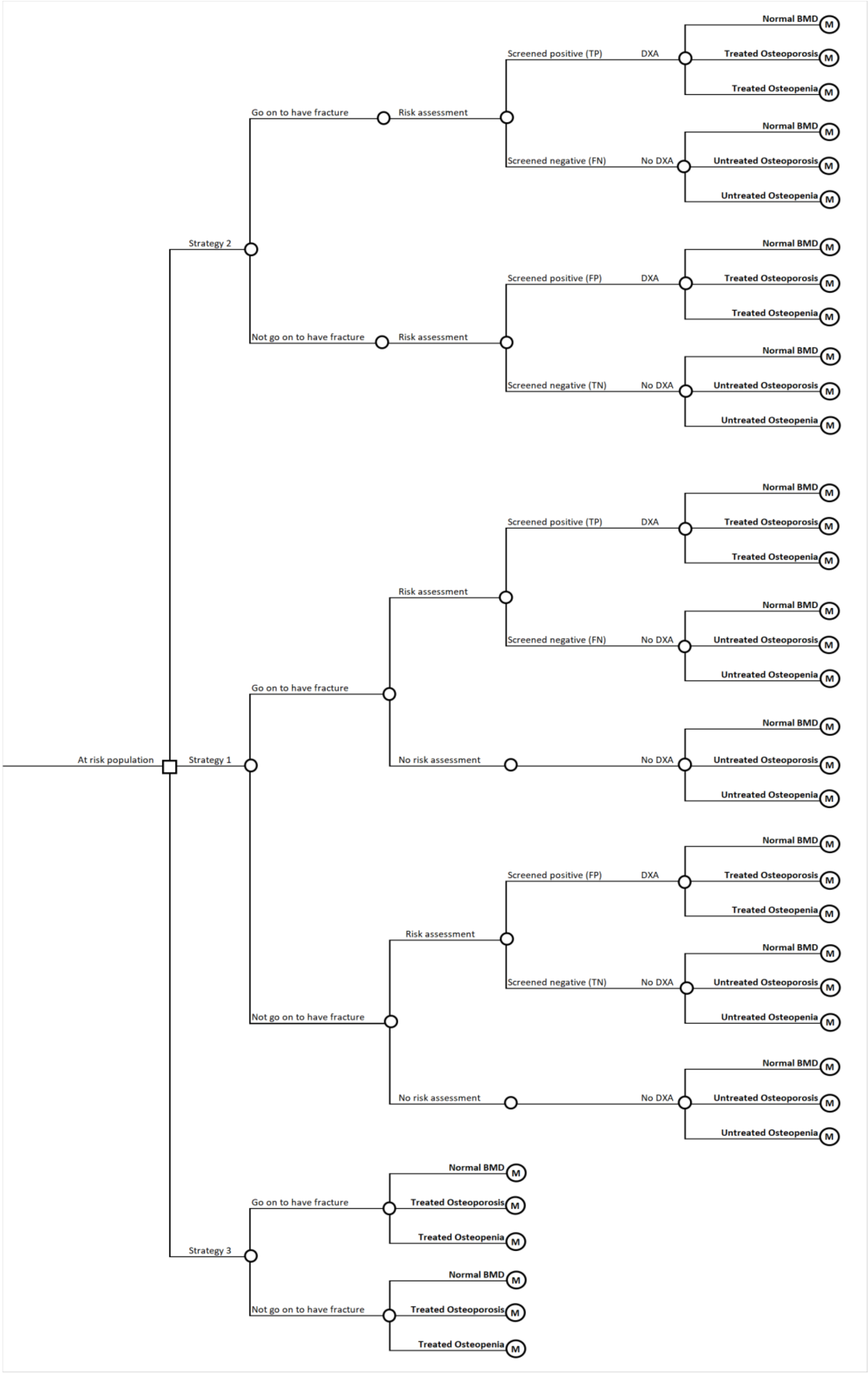


BMD: bone mineral density. DXA: dual-energy X-ray absorptiometry.

**Supplementary Fig. 2. Markov model**

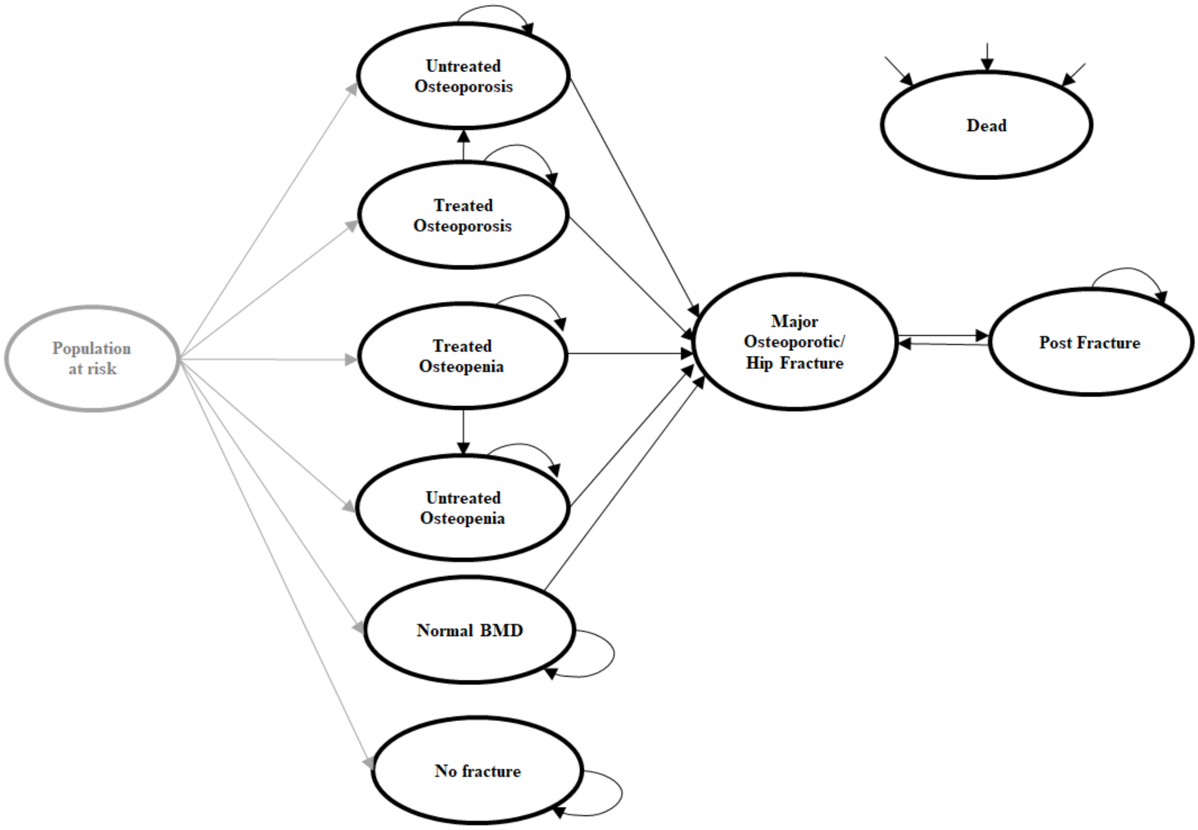


“Population at risk” is not a health state in the Markov model, it represents the movement of people in the decision tree. BMD: bone mineral density.

**Supplementary Fig. 3. Cost-effectiveness analysis of Strategy 2 vs. Strategy 3 among people with intellectual disabilities with major osteoporotic fracture using (a) cost-effectiveness plane and (b) cost-effectiveness acceptability curve.**

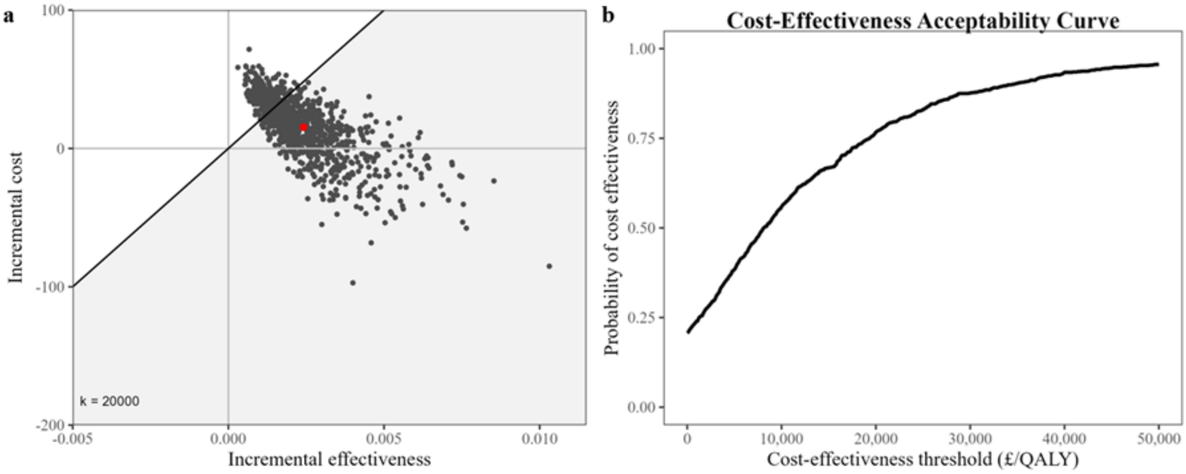

**Supplementary Fig. 4. Cost-effectiveness analysis of Strategy 2 vs. Strategy 3 among people with intellectual disabilities with hip fracture using (a) cost-effectiveness plane and (b) cost-effectiveness acceptability curve.**

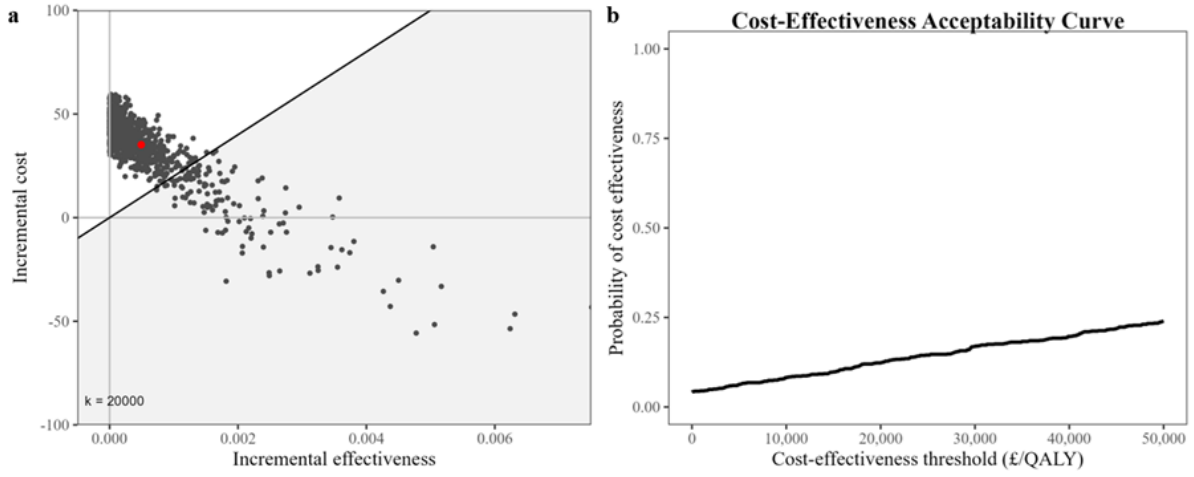
